# COVID-19 impact on Socio-economic and Health Interventions: A Gaps and Peaks analysis using Clustering Approach

**DOI:** 10.1101/2022.01.09.22268991

**Authors:** Hridoy Jyoti Mahanta, G. Narahari Sastry

## Abstract

A quantifiable model to describe the peaks and gaps during the several waves of COVID-19 is generated and applied to the progression of 120 countries. The number of waves encountered and how many more to be encountered is a question which is currently explored by all the scientific communities. In the same quest, an attempt has been made to quantitatively model the peaks and the gaps within them which have been encountered by 120 most affected countries from February 2020 – December 2021. These 120 countries were ranked based on the number of confirmed cases and deaths recorded during this period. This study further cluster these countries based on socio-economic and health interventions to find an association with three dependent features of COVID-19 i.e. number of confirmed cases, deaths and death-infectivity rate. The findings in this study suggests that, every wave had multiple peaks within them and as the number of peaks increased, predicting their growth rate or decline rate turns to be extremely difficult. However, considering the clusters which share the common features even with diverse countries, there is some possibility to predict what might be coming next. This study involves exhaustive analysis of reliable data which are available in open access and marks an important aspect to the COVID-19 research communities.

## 1. Introduction

The evolution of current pandemic from the beginning of 2020 has literally spread to every parts of the world. The impact has been so wide spread and it appears to be hard to comprehend the relationship between the spread of the virus to either socio-economic stratum, medical and health care preparedness or economic power of a nation. While there are disparities in the way the pandemic has impacted the nations at different stages, a bird’s eye view reveal that after the two years, virtually no country could escape from the dreadful consequences of the pandemic in one way or the other.

Epidemics and pandemics are not new disasters which the world has never seen before. As mentioned by Dobson and Carper [1], as human race shifted from being hunter-gatherers to agrarians, it has favoured the spread of various infectious diseases. With the increase in trade among communities, cities, territories and so on, the emergence and spread of infectious diseases has grown exponentially. This has actually led to high risk of outbreaks, epidemics and even pandemics [2]. Plague, which was one of the first kinds of pandemics the human race has ever seen, appeared in the 5^th^ century for first time and eventually reappeared in the 13^th^ century causing massive human deaths (30% of European population) and hence was known popularly as Black Death [3]. The world was again collapsed in the beginning of 19^th^ century when it encountered the Cholera pandemic. During the entire 19^th^ century, Cholera was encountered more than 5 times by the human race in almost all parts of the world causing death to millions of human lives. As reported by WHO, Cholera still affects 1.3 – 4.0 million of the world population every causing death of almost 100 thousand lives [4, 5]. The Spanish flu and its counterparts of the 20^th^ century which were caused by Influenza also emerged as pandemic killing almost 50 million peoples [6, 7]. The zoonotic transmission of pathogens from animals to humans is a key mechanism which leads to emergence of infections and has bothered humans throughout history [8]. This was clearly seen in the 21^st^ century with the outbreaks like Severe acute respiratory syndrome (SARS), Middle East respiratory syndrome (MERS) in which Bats, dromedary Camels, palm civets were the major vectors [9, 10]. Other factors which have influenced the transmission of pathogens to a large extent include climate change, geographic locations and excessive population growth [11-14].

In December 2019, a group of patients in the Wuhan city of China were reported with a typical pneumonia caused by a new coronavirus SARS-CoV-2 also referred as COVID-19 [15]. Bats and Pangolins were predicted to be the animal hosts for the outbreaks, however the intermediate host still remains unidentified [16, 17]. Within few months of its inception, COVID-19 spread all over the world leading to more than 74 million contaminations and 1.6 million deaths in December 2020 [18]. This rose to 298 million contaminations and 54 million deaths in December 2021 [19] and with more than 2 million cases in every 24 hours, almost ≈ 36% of the world population got infected with this virus. Apart from claiming the lives, COVID-19 has also shattered the health care systems [20-22] and economy [23, 24] of most of the developed countries across the world. Many mathematical models have been developed by researchers to look at the impact of COVID-19 on different factors like prediction and propagation, trends, quarantine times, vaccinations, socio-economic and technological interventions, sustainability development goals etc. [25-31]. Studies were also carried out on modelling the association of COVID-19 infections with specific issues human habits like smoking, alcohol consumption and demographic factors like population density, climate change problems etc. [32-35]; however with the consistent increase in the rate of infections and new SARS-CoV-2 variants [36] showing up, it is difficult to predict what is going to happen next.

The impact of Artificial Intelligence (AI) and Machine Learning (ML) in developing predictive models is seen across all the fields of scientific discoveries and COVID-19 research is no different. A large number of predictive models have used ML to study the propagation, prevalence as well as control against COVID-19 pandemic [37-45] which shows that these approaches can extensively contribute in identifying the risk of infections, screening and diagnostics, speed up drug discovery/development, predicting risk of new pandemics etc. An effective method to understand the deeper insights of what is going to happen next with this pandemic is to pursue the past data in details. In this study, an attempt has been made to study COVID-19 related features like infectivity, deaths and death-infectivity rate and all possible cross-correlation factors to understand the hidden associations between them. The analysis involves a mathematical model built with mean absolute deviation (MAD) to identify the peaks and their intensities which were visible during all the waves encountered by a group of countries. This group consisted of 120 countries which were ranked highest in number of infections (or confirmed cases) and deaths in July 2021 and were studied for a period of 23 months (February 2020 to December 2021). Along with the peaks and their intensities, the gaps (duration between two peaks) were also calculated to study the trends in which these gaps varied with the number of peaks. Further, the skewed distribution was calculated using the concept of area under the curve (AUC) to identify the rate of growth as well as decline to the ground for each peak. Finally, 23 independent features related to socio-economic, environment and health factors were correlated with the COVID-19 related features and the countries were clustered using k-means clustering. Then intra-cluster analysis was performed to identify the how closely these 23 features were associated with the number and intensities of the COVID-19 peaks to give key insights in identifying the possibilities of new waves and peaks in near future. What differentiates this study from others is that it involves a large number of countries and analysis with COVID-19 data right from inception till current period. This huge data has been translated into peaks and gaps for interpretation and converged in finding the association with some key features which can be potential indicators for future outbreaks.

## 2. Materials and Methods

### 2.1 Data collection and preparation

The COVID-19 data for 120 highly affected countries ranked by the number of infections (or confirmed cases) and confirmed deaths in period July 2021 have been considered for this study. The baseline for selecting a country for this study was that it should have a minimum of 1000 cases/day and 100 deaths/day to consider it as one of the highly affected countries. The data has been extracted from *https://worldometer.info* [46] and *https://ourworldindata.org* [47]. The primary data which have been considered for this study from these sources are number of confirmed cases with their times lines and confirmed deaths. Apart from these, 23 feature variables based on four indicators – COVID-19 indicators, socio-economic indicators, environmental performance indicators and health indicators were extracted from different sources and considered in this study as shown in **Table 1**. The COVID-19 indicators comprises of excess death, skewed distribution of peaks and mean of gaps between the COVID-19 peaks for each country. The excess death feature was taken from *https://ourworldindata.org* [47] but the other two indicators of COVID-19 were computed separately. The socio-economic indicators were the parameters like population density, GDP, average life expectancy, median age, percentage of senior citizens, per capita alcohol consumptions, and percentage of smokers (male and female). These features were taken from the world global health observatory [48]. The environmental performance indicators featured the air quality, unsafe drinking water, average annual temperature, air pollution and environmental performance index (EPI) extracted from *https://epi.yale.edu* [49]. In this study, we have also examined the health indicators with country wise features like BMI in adults, mortality rate due to non-communicable diseases and unsafe water, sensitisation and health (WASH) along with the probability of deaths by different diseases like Cancer, Diabetes, lower respiratory disease and COPD which were also collected from world global health observatory [48].

**Table 1.**
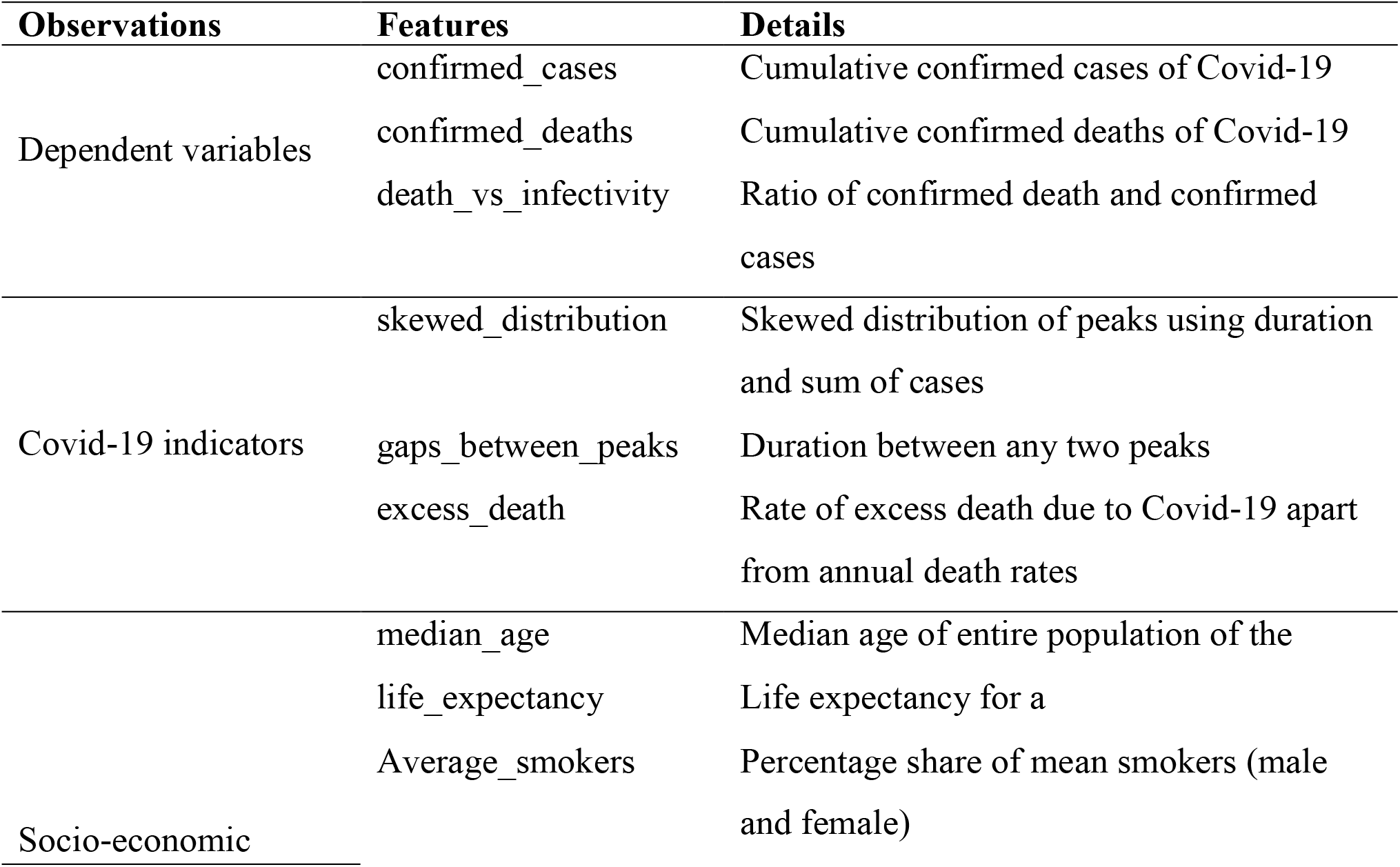
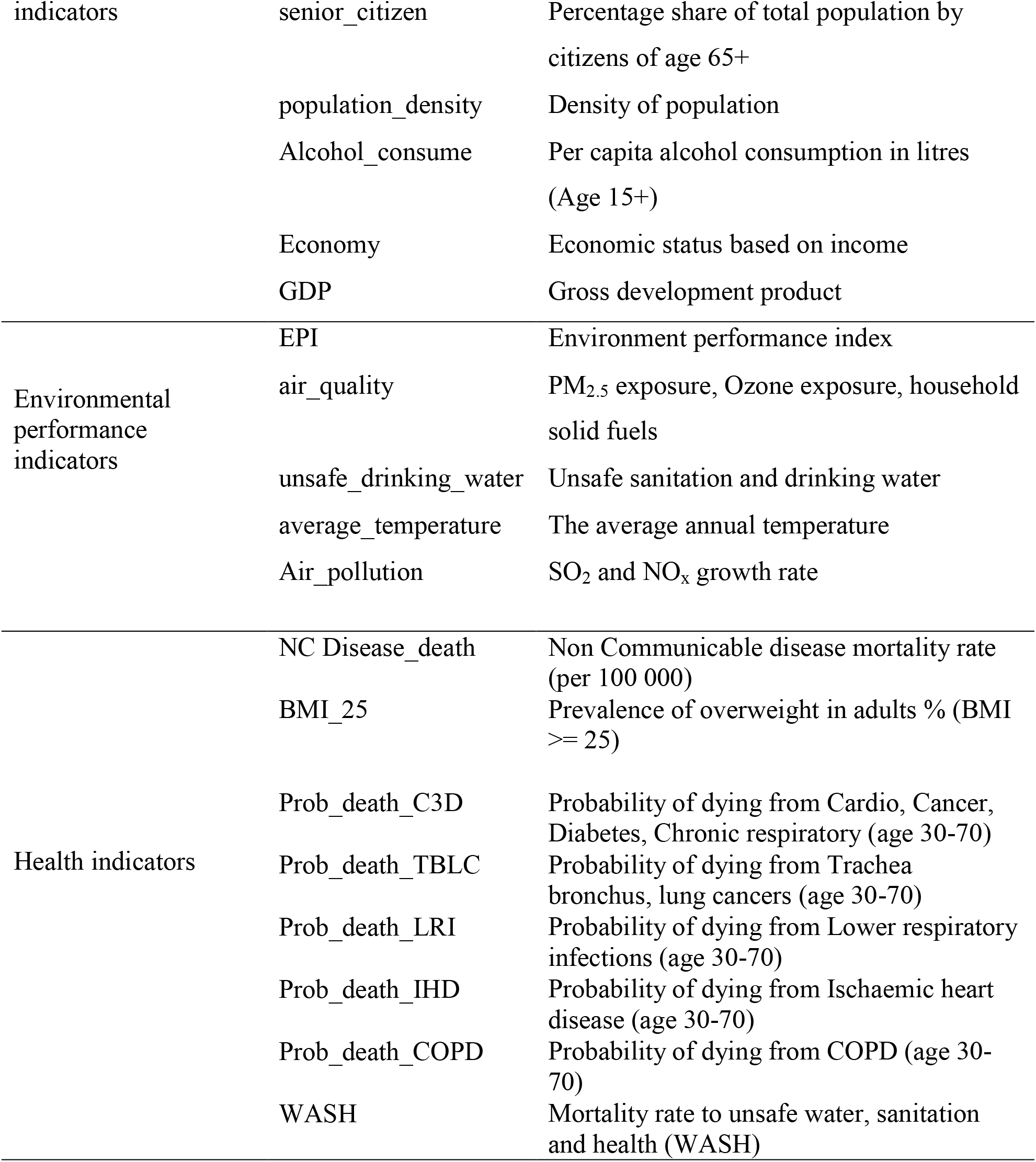
Detailed overview of the indicators considered for clustering 120 countries ranked top based on confirmed COVID-19 cases and deaths.

As the data collected for this study was widely diverse and had large variance, they were standardized to remove the mean value of each feature and then divide it by the standard deviation using the formula,

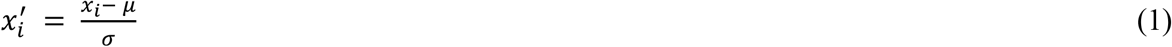

Where, 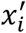 is the standardized value against *x*_*i*,_ which is a feature value of the set for X with *μ* and *σ* being the mean and standard deviation respectively.

### 2.2 Identifying the peaks and gaps

Whereas most of the mathematical models in COVID-19 research have used cumulative curves in their studies, in this work we have explored the epidemic curve or waves with respect to number of confirmed cases which a country has recorded for a period of 23 months (February 2020 – December 2021). One way to identify the waves is to look at the R-factor (reproduction number) which is the average number of people infected by one infectious person. If the value of R is significantly > 1 for a sustained period, that time period can be considered as an upward period (rise), and inversely, if R is significantly smaller than 1 for a sustained period, then that period is a downward period (fall) leading into formation of a wave. Another way is to calculate the mean absolute deviations (MAD) [50] over a fixed period of time which helps to identify the variation of in the number of cases during that specific period. This study is based on the hypothesis that if the MAD of daily variation is less than 10% for a period of 40 days, then it may be a gap or plateau. The MAD values depict the peaks with respect to the time whose intensity represented the number of confirmed cases during that period. Between any two peaks, the difference or time gap has been used to represent the gaps between them as shown in Figure 1.

**Figure 1.**
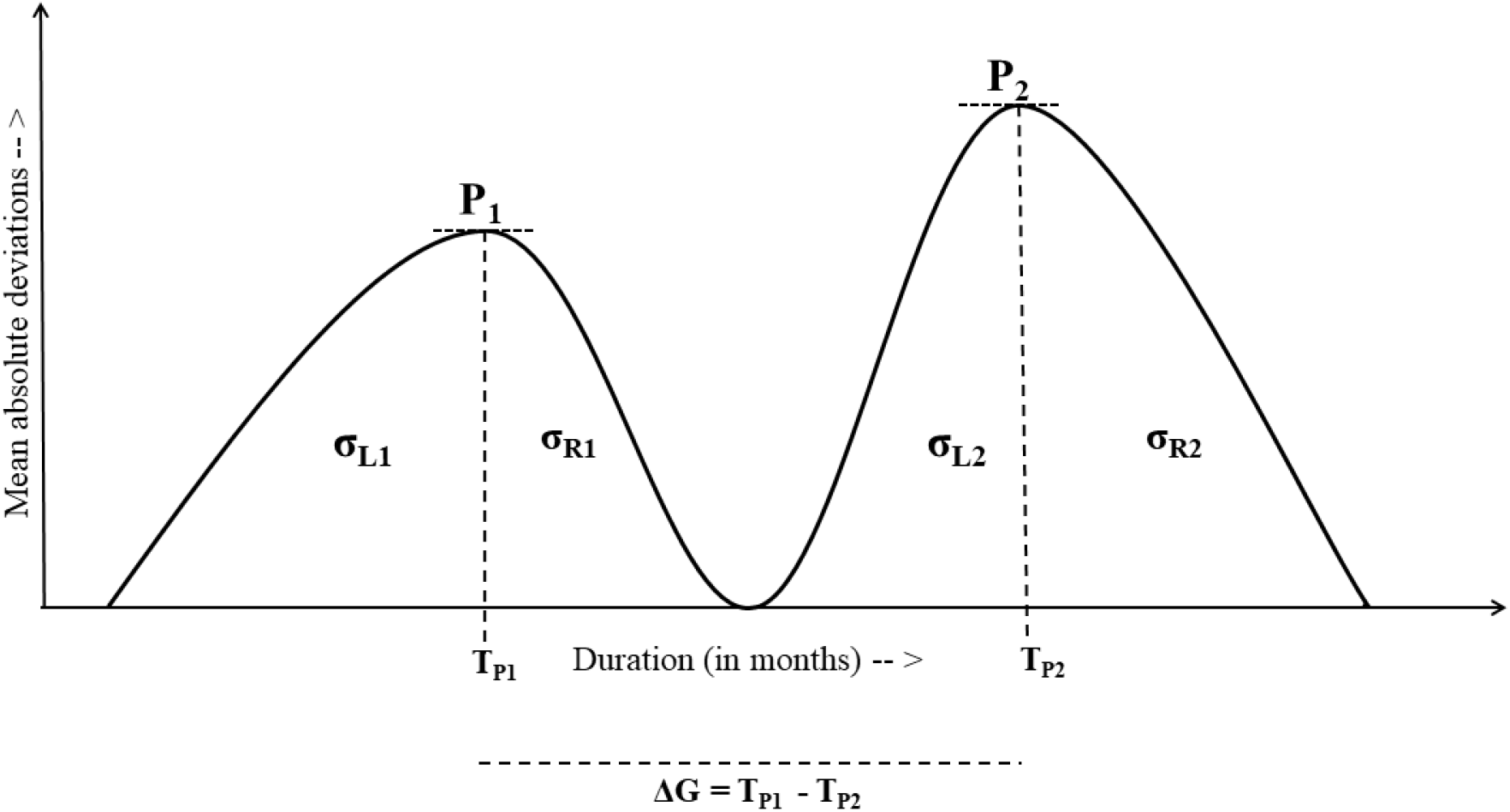
Schematic representation of different concepts used in this study: P_1_ and P_2_ are the peaks, T_P1_ and T_P2_ are the points where peaks appeared, σ_L_ and σ_R_ are the skewed distributions with respect to each peak and ΔG is the gap between the peaks.

An advantage of using the MAD based mathematical model for identifying the COVID-19 peaks is that also leads to find the waves where the peaks lies. Basically, during a wave, the spread of infections increases drastically which means the number of new cases during this period is substantially high till it reached a peak point after which the cases comes down till it reaches a valley or gets neutralized. A wave can have multiple peaks out which one will be the highest among all. For identifying the peaks, month-wise MAD was computed for each country from February, 2020 till December, 2021. The computation of MAD was done by,

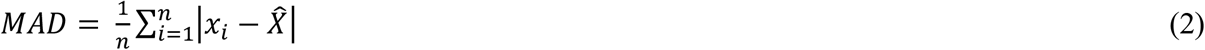

Where *x* is the number of daily cases and 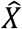 is the mean during the period *n* (number of days).

If *P*_*i*_ is an identified peak then using MAD, the necessary conditions it satisfies are,

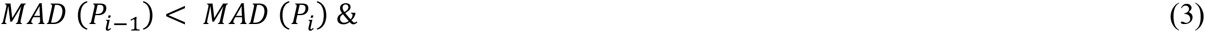

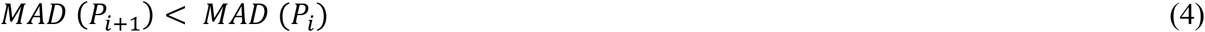

However, between any two identified peaks there may also be a valley between them such that if *v*_*i*_ is a valley between two peaks at a point *p*_*i*_ than at *v*_*i*_

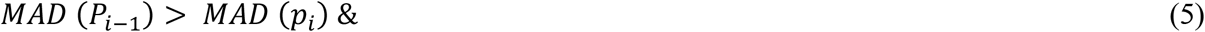

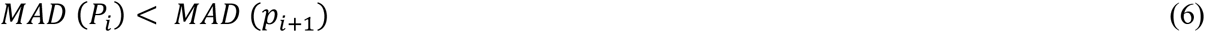

As a wave can have multiple peaks, the gaps among them are also important factors to calculate for justifying our hypothesis. Mathematically, if any two peaks *P*_*i*_ and *P*_*i+1*_ have occurred in the time period *T*_*Pi*_ and *T*_*Pi+1*_ respectively then the gap (ΔG) between the peaks will be,

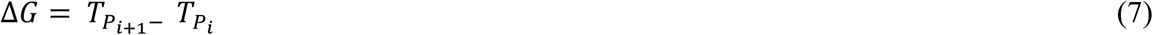

Apart from the gaps, the width or duration of peaks was also computed to find out the period for which each peak existed and identify the period of rise and period of decline or reaching the valley. If *L*_*i*_ represents the start of the *i*^*th*^ peak and *L*_*i+1*_ is the point of end for that peak then the width or duration (δ) of the peak can be calculated by,

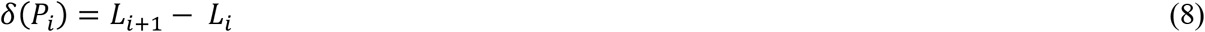

### 2.3 Analysis of the skewed distribution of peaks

In order to analyse the steepness of increase and decline of the COVID-19 peaks during the waves, we have computed the area under the curve for the left and right regions under each peak. The left area (AUC_L_) and right area (AUC_R_) under the peak gives an absolute measure on which part of a wave the number of cases was higher relative to its duration which results into a plateau. To compute AUC, relative areas on left and right side of the peak (α_L_ and α_R_ respectively) were computed in such a way that for peak P_1_,

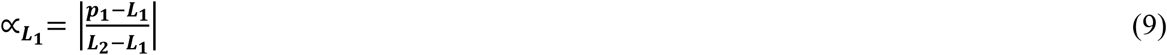

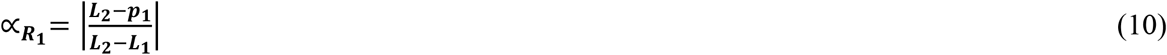

Where, *p*_*1*_ was the period at which peak was arrived and *L*_*1*_ and *L*_*2*_ are the starting and end of the peak P_1_. To generalize the equations (9) and (10) for any peak *P*_*i*_ we can hence have,

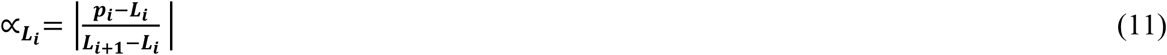

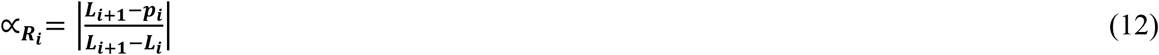

Then the summation (σ) of all the new cases (*n*) during these period are computed for both left and right areas of the peak such that,

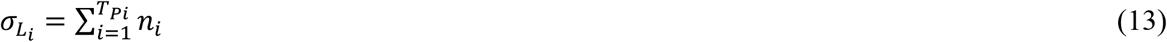

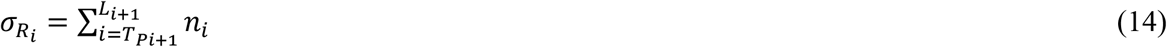

Finally, the ratio of area under the curves in left and right are computed such that,

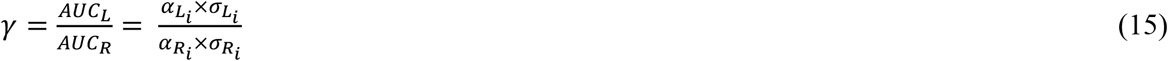

The ratio (γ) could determine whether the peaks in the waves had steep increase or decline with the fact that if γ > 1 then the left area under the peak is greater than right area resulting slow increase of cases or infectivity and if γ < 1 then right area under the peak is greater than left area depicting gradual decline in the cases or infectivity.

### 2.4 Finding the correlation of the features

We have calculated the correlation of the feature variables with the COVID-19 related factors i.e. death-infectivity rate, confirmed cases and confirmed deaths. The Spearman’s rank correlation coefficient [51, 52] determines the strength and association between two variables having monotonic relationships and can be derived by,

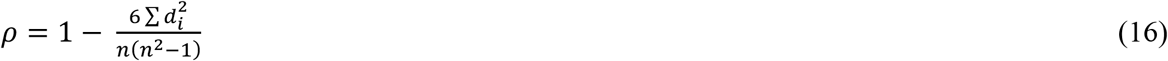

Where, *d*_*i*_ is the difference in paired ranks and *n* is the total number of case. The value of the coefficient lies between +1 to -1 where, +1 signifies perfect positive correlation between the ranks, -1 signifies perfect negative correlation and 0 signifying no correlation at all. Equation (16) works when there is no tie among the ranks, however if there is a tie than the correlation coefficient with paired score *i* is determined by,

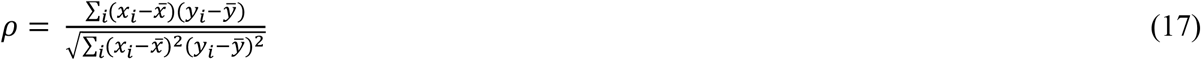

### 2.5 Clustering the countries

Based on the 23 feature variables and the three COVID-19 related factors, we have clustered the countries into multiple clusters. K-means clustering [53, 54] algorithm which facilitates centroid based partitional clustering was used to form 4 clusters out of the 120 countries. The reason behind using K-means clustering was its simplicity and effectivity in forming clusters with high intra-cluster and low inter-cluster similarity. The number of cluster centres was predefined using the Elbow method. In every iteration, the mean cluster centres are updated depending on the newly assigned data points having minimum distances from the cluster centres.

The feature variables of each country were assigned membership for defining the initial *k* clusters. Based on the minimum distance from the cluster centres, the data point associated with each country was assigned to one of the 4 clusters and eventually the cluster means were updated. This resulted into feature variables with similar properties led to group the respective countries into the same cluster. These clustering not only benefitted to derive relationships between the factors leading to cluster membership but also understand the different aspects leading to more number of confirmed cases, deaths as well as the death-infectivity rate.

### 2.6 Performance analysis of clustering

In order to measure the performance of clustering in this study we have computed the inertia, Silhouette coefficient [55, 56], Calinski-Harabasz index and Davis-Bouldin index [57]. As the ground truth in clustering is unknown, hence these parameters will indicate how accurately the clusters were formed. While forming the clusters using k-means clustering, inertia depicts the distance between each single data point and its respective centroid, squaring this distance, and summing these squares across one cluster. Inertia with *k* clusters for a set of observations *S* with *x*_*i*_ being a data point and *μ*_*i*_ as the mean of all data points can be calculated by,

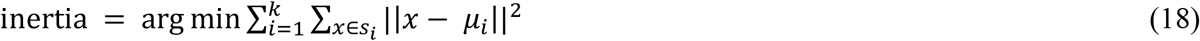

An ideal model has less number of clusters and smaller inertia value however, with the increased number of clusters, the inertia may decrease leading to tradeoff.

The Silhouette Coefficient is also used to analyze the clustering model when ground truth is unknown. This coefficient is defined as the ratio of mean distance between a sample and all other points within the same class and mean distance between a sample and all other points in the next nearest cluster. The Silhouette score can is calculated by,

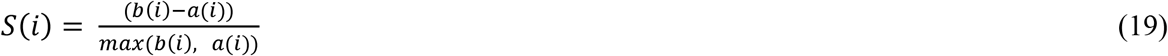

Where, *a(i)* and *b(i)* represents intra-cluster and inter-cluster distances for the *i*^*th*^ point from all other points. The Silhouette scores lies between the range from -1 to +1 which depicts the fact that higher is Silhouette coefficient score, better are the clusters defined.

The third parameter calculated for evaluating the performance of clustering model is the Calinski-Harabasz (CH) Index. It is the ratio of the sum of between-clusters dispersion and of within-cluster dispersion for all clusters (where dispersion is defined as the sum of distances squared). It is also known as the variance ratio criterion and can be computed by,

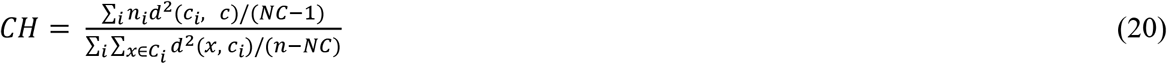

Where, *n* is the number of data points in the dataset, *d* is the distance of the point *x* from the center of the *i*^*th*^ cluster *C*_*i*_, and *NC* is the number of clusters. A higher Calinski-Harabasz score relates to a model with better defined clusters.

The last parameter used for performance evaluation is the Davies-Bouldin Index (DB) which signifies the average ‘similarity’ between clusters, where the similarity is a measure that compares the distance between clusters with the size of the clusters themselves. It is computed by,

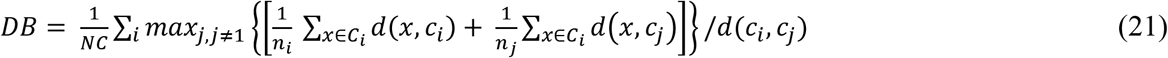

The lowest possible score of Davis-Bouldin index is 0 and maximum is 1. With values closer to zero indicates a better partition or better separation between the clusters.

## 3. Results and Discussions

### 3.1 Identifying the peaks and gaps

The mean absolute deviation was calculated with respect to the number of confirmed cases for each month from the period where the number of cases were more than 1000 and deaths more than 100 per day. From the analysis it was seen that the 117 out of 120 countries had at least 1 peak during the pandemic with Netherlands having maximum of 7 peaks followed by United States, Chile and Pakistan with 6 peaks each. It is worth mentioning that the number of peaks was not in aligning with the number of waves i.e. a wave was seen to have more than one peaks in some cases. Hence, in order to identify the waves for each country we underwent a visual inspection at *https://worldometer.org* [46]. The findings of the waves and peaks for each country have been shown in Figure 2. During the visual inspection of the waves, the pattern of the waves was also monitored to understand which wave had the highest intensity among all the waves for a particular country. This observation helped to find out that maximum number of countries (81 out of 120) had a higher second wave than all other waves which was followed by higher first wave for 24 countries and higher third wave for 14 countries. South Africa was the only country found to have the fourth wave highest among all the waves. The emergence of the new variant with highest doubling time justifies this fact. **Supplementary Table 3** shows the countries with their respective waves where the intensity was highest among all.

**Figure 2.**
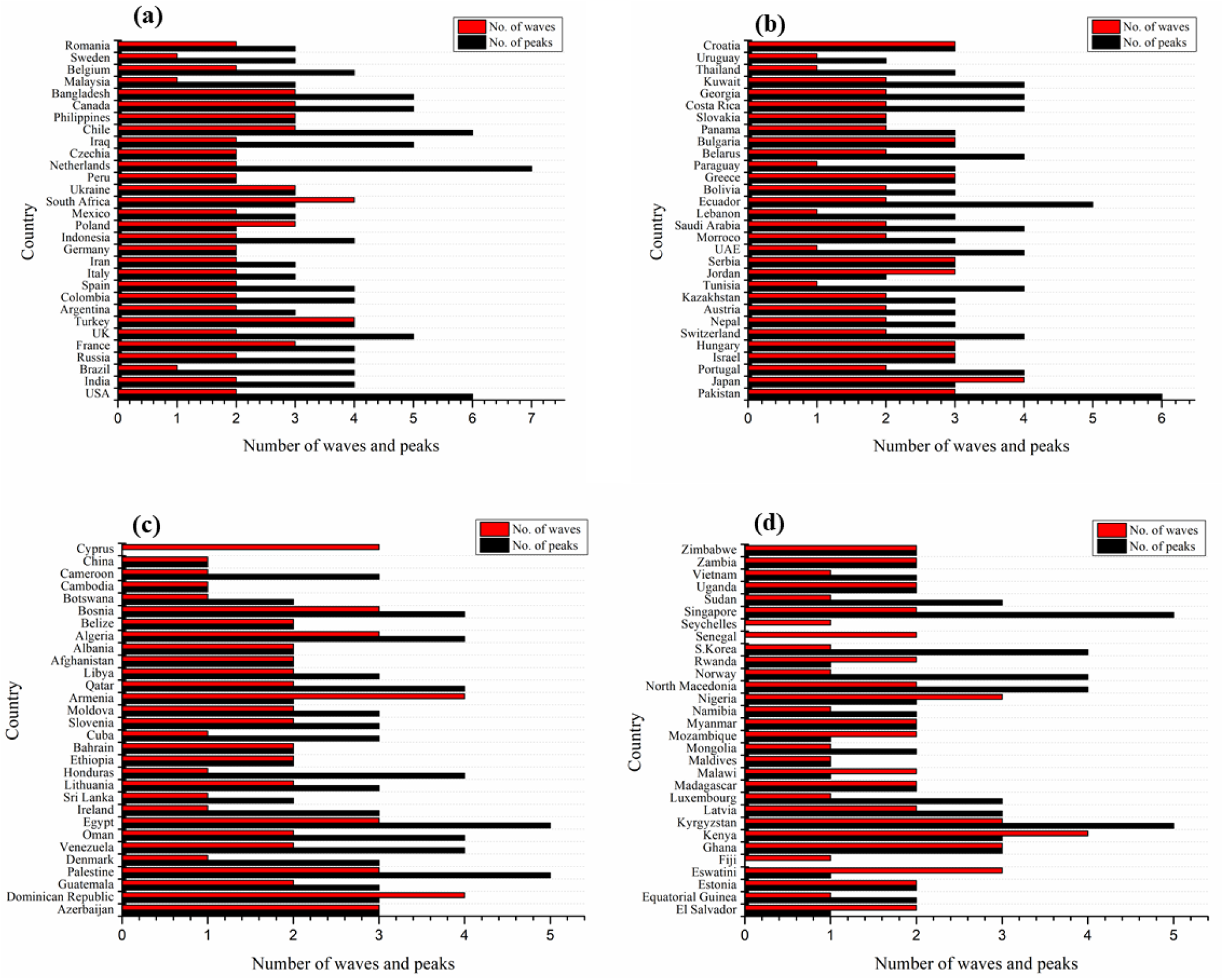
Number of waves and peaks for top 120 countries having highest number of Covid-19 cases and deaths calculated in this study with mean absolute deviation. Each individual figure (a-d) consists of 30 countries each.

Along with the waves and peaks within them, the gaps between the peaks were also measured to study the variation in time periods in formation and collapse of these peaks. For the first and second peaks, it was observed that the maximum duration was 304 days for Equatorial Guinea whereas the minimum was 59 days for Algeria. On average considering all the 120 countries, an average of 138 days gap was there between the first and the second peak. Between the second and third peak, it was observed that Zimbabwe had the highest gap of 251 days and Norway had lowest gap of 59 days and on average it took around 130 days to arrive at the third peaks. For the fourth peak, Zimbabwe again had the highest gap of 212 days and Chile as least with 59 days making an average of 113 days between these peaks. It worth mentioning that not all countries had 3 or more peaks, the analysis has considered only those countries that have seen these multiple peaks. As we can see, the maximum and average gaps between the peaks go on decreasing as the number of peaks increased. These analysis results have been shown in details in **Supplementary Table 4**.

### 3.2 Analysis of skewed distributions

The skewed distribution analysis was performed for each peak for all the countries. This analysis depicts two primary facts – a) whether the growth of infectivity (or confirmed cases) was steady till it reached the peak and b) whether the decline of cases was faster to reach a plateau or neutralized level and vice-versa. In this regard, three aspects were investigated for every possible distribution of the skews. These were, *left bound, right bound* and *mixed distribution*. Left bound interpreted that the growth in the number of cases was steady and the duration to reach the peak was considerably higher than its decline rate. Similarly, right bound indicated that the rate of decline to cases to reach a neutralize level or start another peak was higher and longer than the growth rate. Finally, there were cases where for countries with multiple peaks, some peaks were left bound where as others were right bound, this gave have been considered as mixed distribution in this study. The analysis results shows that out of 120 countries, 30 countries were left bound which included mostly the European countries where as there were 11 countries which were right bound like Czechia, Nepal, Greece, Slovakia, Sri Lanka etc. In both cases the maximum number of peaks for these countries was 4. A large number of countries (74 countries) were found to a have mixed distribution with the COVID-19 peaks. These included those countries which have seen highest number of peaks during the period i.e. Netherlands (7 peaks), USA, Chile, Pakistan (6 peaks) and so on. The distribution of the countries based on this analysis has been shown in **Figure 3 (a - c)** including other factors like number of waves and peaks encountered. The detailed results of this skewed distribution has been illustrated in **Supplementary Table 6**

**Figure 3.**
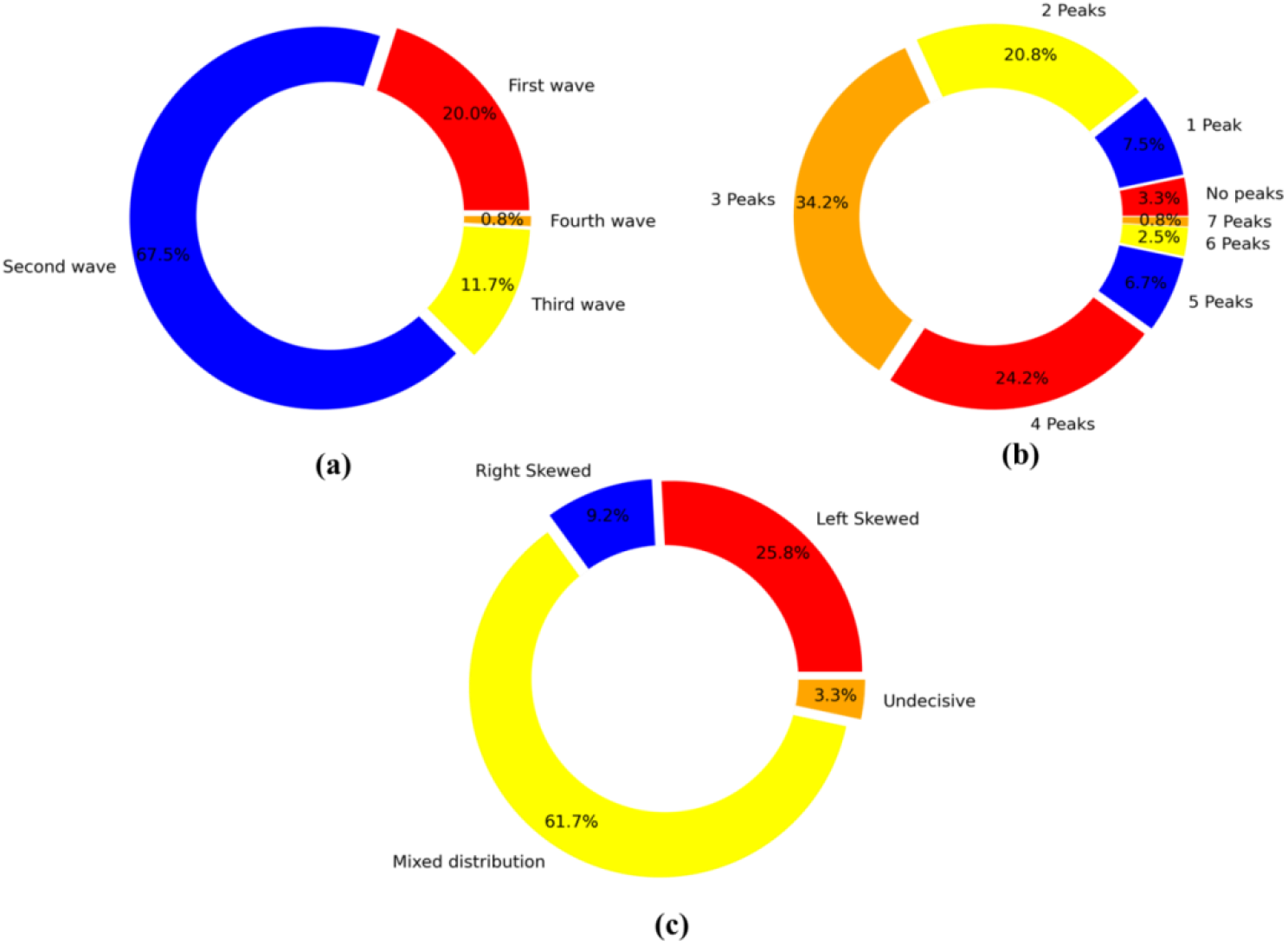
Distribution of the 120 countries considered in this study based on three factors **(a)** Number of waves encountered **(b)** Number of peaks and **(c)** Skewed distribution of the peaks

### 3.3 Correlations with feature variables

The Spearman’s correlation of the 23 feature variables with the COVID-19 related features depicts a monotonic association among them. From Table 2, it can be interpreted that with respect to death-infectivity rate, the correlation scores are very weak and there are many features having negative correlations. Moderate correlation can be seen with excess death rate (**0.241**) which is above the normal death rate of a country and the prime reason for this is definitely the COVID-19 pandemic. With all other features, the correlation score is very close to zero and hence is difficult to derive strong conclusion.

**Table 2.**
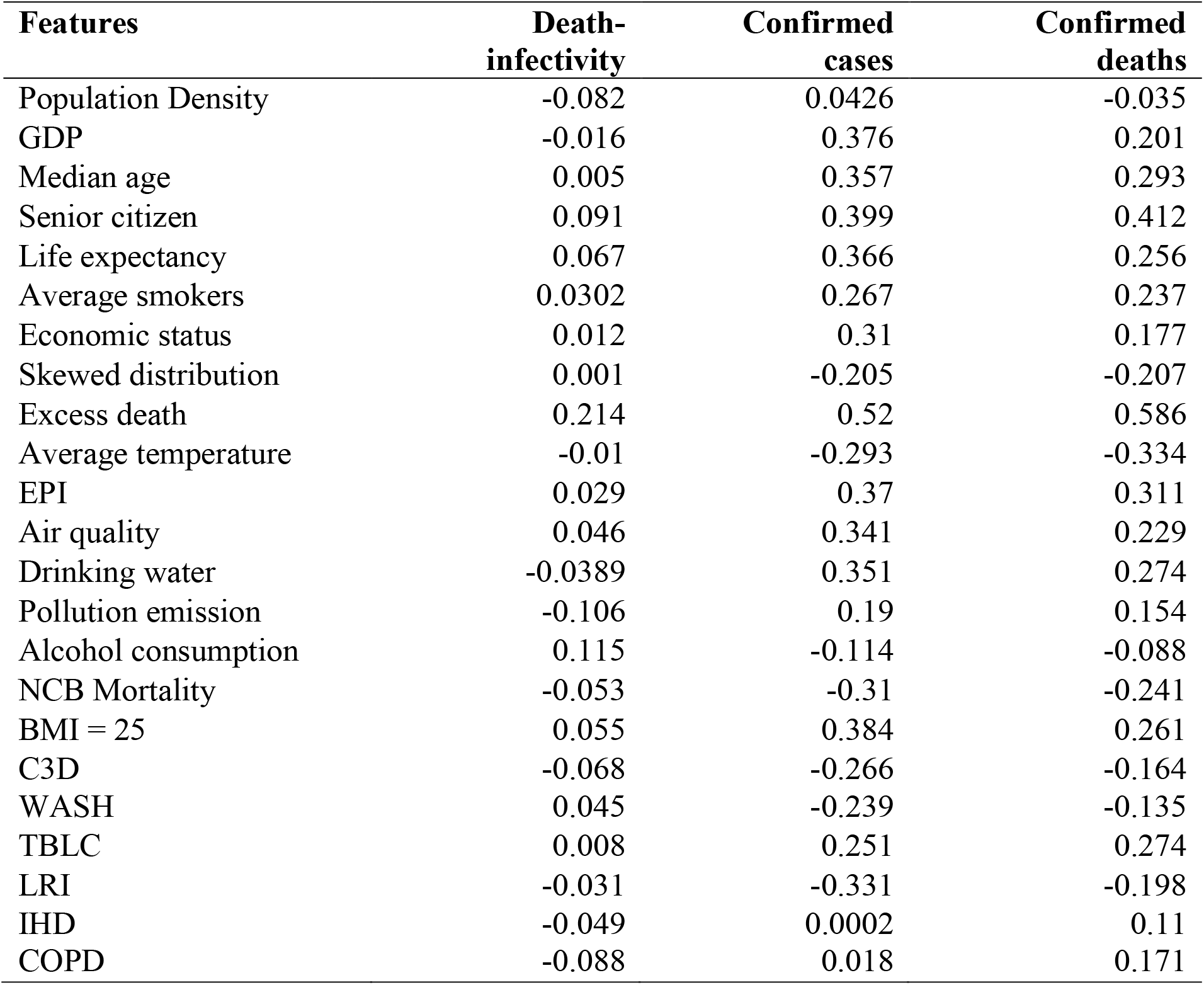
Spearman’s correlation coefficient for 23 features with respect to death-infectivity, confirmed cases and confirmed deaths

| Features | Death-infectivity | Confirmed cases | Confirmed deaths |
| --- | --- | --- | --- |
| Population Density | -0.082 | 0.0426 | -0.035 |
| GDP | -0.016 | 0.376 | 0.201 |
| Median age | 0.005 | 0.357 | 0.293 |
| Senior citizen | 0.091 | 0.399 | 0.412 |
| Life expectancy | 0.067 | 0.366 | 0.256 |
| Average smokers | 0.0302 | 0.267 | 0.237 |
| Economic status | 0.012 | 0.31 | 0.177 |
| Skewed distribution | 0.001 | -0.205 | -0.207 |
| Excess death | 0.214 | 0.52 | 0.586 |
| Average temperature | -0.01 | -0.293 | -0.334 |
| EPI | 0.029 | 0.37 | 0.311 |
| Air quality | 0.046 | 0.341 | 0.229 |
| Drinking water | -0.0389 | 0.351 | 0.274 |
| Pollution emission | -0.106 | 0.19 | 0.154 |
| Alcohol consumption | 0.115 | -0.114 | -0.088 |
| NCB Mortality | -0.053 | -0.31 | -0.241 |
| BMI = 25 | 0.055 | 0.384 | 0.261 |
| C3D | -0.068 | -0.266 | -0.164 |
| WASH | 0.045 | -0.239 | -0.135 |
| TBLC | 0.008 | 0.251 | 0.274 |
| LRI | -0.031 | -0.331 | -0.198 |
| IHD | -0.049 | 0.0002 | 0.11 |
| COPD | -0.088 | 0.018 | 0.171 |

However, the results are better in with respect to confirmed cases and deaths. The socio-economic feature variables have moderate correlations with confirmed cases with senior citizen (**0.399**) being the highest. Similarly, the features related to environmental indicators also shows positive correlations with confirmed cases with drinking water (**0.351**) having the highest association. Whereas among the heath indicators, apart from the BMI variable (**0.384**) and probability of death due to TB and lung cancer (**0.251**), no other features shows positive correlation with the COVID-19 cases. A moderate negative correlation can be seen in case of probability of death due to diseases like lower respiratory infections, Cancer, Diabetes etc.

In case of the confirmed deaths, we can interpret similar associations of the feature variables like confirmed cases. Moderate correlation can be seen in with the socio-economic features with senior citizen having highest association (**0.412**). The features associated with the environmental indicators also had a moderate positive correlation with EPI being the highest (**0.311**). In case of health indicators, other than BMI and probability of death due to TB and lung cancer, all other feature variables had negative correlation with the confirmed deaths.

Interestingly, it could be seen that the average temperature had a negative correlation with all the three parameters considered in this study. Also, excess death had high association with each parameters proving that the case of excess death is only due to COVID-19 pandemic. Further, an observation at the skewed distribution feature, which shows the rate of rise or decline of cases, has mainly negative correlation depicting the fact that if the growth of cases is slow, the number of confirmed cases/deaths will be less and vice versa. This can relate to the actual scenario that as the number of cases increases slowly, there is number of patients who recovers too during that period. Also, with fewer numbers of cases, there is a chance that all the patients will get available medical facilities and so both confirmed cases and deaths will be less. Whereas, on other hand if there is a fast growth in number of cases, the number of deaths will be more due to lack of hospitalization or getting necessary medical facilities. Figure 4 depicts a graphical representation showing the correlations of these 23 independent features with the COVID-19 features.

**Figure 4.**
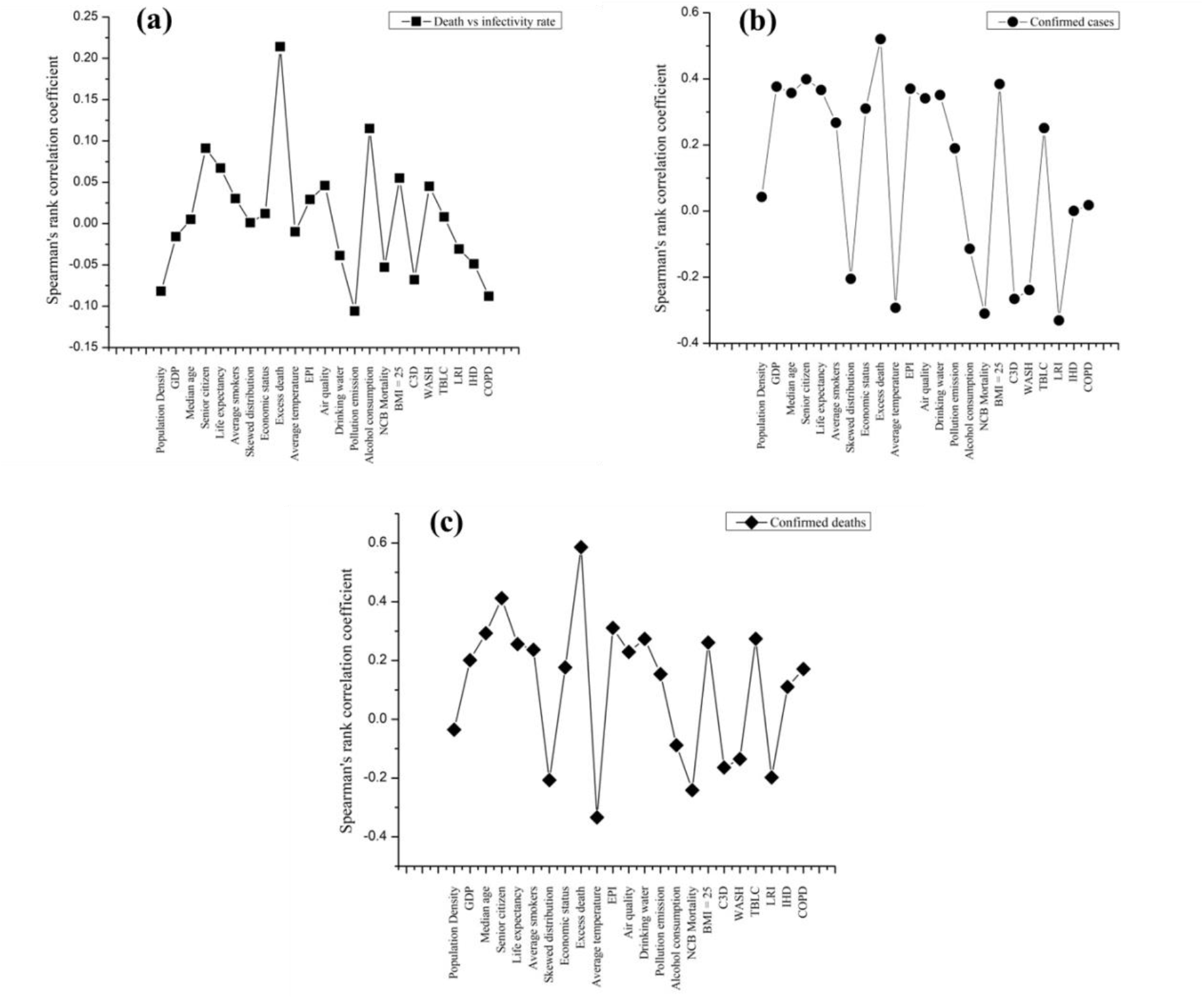
Spearman’s correlation coefficient for 23 indicators against (a) death-infectivity rate (b) confirmed cases and (c) confirmed deaths

### 3.4 Clusters analysis

Four clusters were formed while associating COVID-19 death-infectivity rate, confirmed cases and confirmed deaths with 23 features related to socio-economic factors, environment and health. The optimal number of clusters was predefined using Elbow method as can be seen in Figure 5. The cluster means of these features with respect to the COVID-19 features have been illustrated in Table 3. Among these four clusters, the first cluster comprised of 28 countries as in Table 4 (a) which has second highest cluster mean of COVID-19 death-infectivity rate (0.207), last in number of confirmed cases (−0.158) and third highest in number of confirmed deaths (−0.100). This cluster included countries like Sudan, Afghanistan, Myanmar, Malawi which had a high rate of death-infectivity due to COVID-19. The average death-infectivity rate in this cluster was 2.28% as per the data available in [46]. The high cluster mean of health indicators like probability of death due to heart disease, lung cancer were very high in these countries. Most of the countries in this cluster are low and lower-middle income countries with very low GDP and poor environmental factors like high air pollution, poor water sanitation and very less annual rainfall.

**Figure 5.**
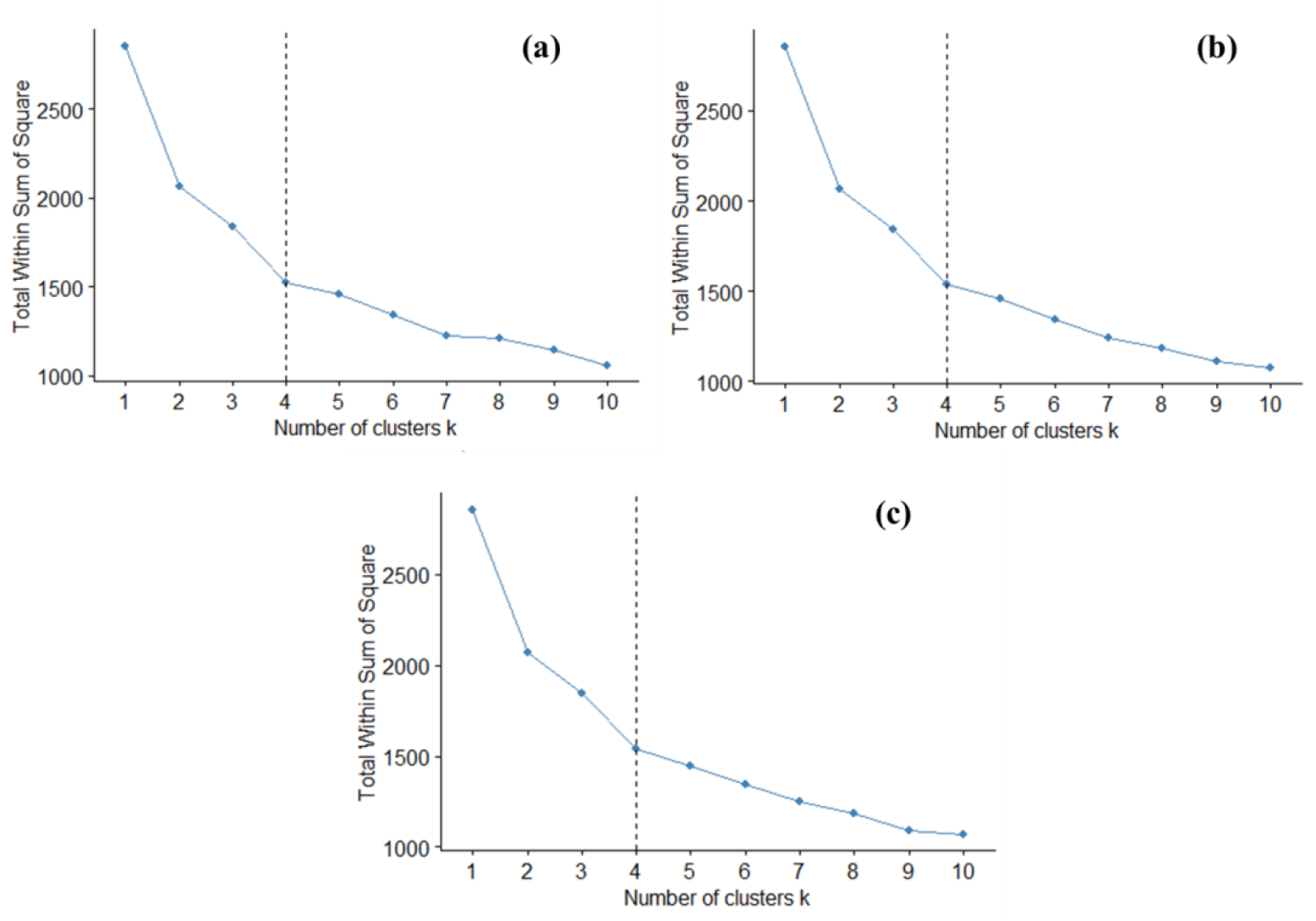
Choosing the optimal value of *k* using elbow method for **(a)** death-infectivity rate **(b)** number of confirmed cases **(c)** number of confirmed deaths

**Table 3.** Clusters mean of 23 feature variables with respect to death-infectivity rate, confirmed cases and confirmed deaths from Covid-19.

| **Features** | **Cluster 1** | **Cluster 2** | **Cluster 3** | **Cluster 4** |
| --- | --- | --- | --- | --- |
| Death-infectivity rate | 0.207 | 0.131 | 0.227 | -0.585 |
| Number of confirmed cases | -0.158 | -0.059 | -0.075 | 0.286 |
| Number of confirmed deaths | -0.100 | 0.041 | -0.130 | 0.141 |
| ***Socio economic indicators*** |  |  |  |  |
| Economy | 0.270 | -0.125 | -1.132 | 1.122 |
| Life expectancy | 0.180 | 0.107 | -1.410 | 1.109 |
| Senior citizen | 0.856 | -0.589 | -0.940 | 1.202 |
| Median age | 0.922 | -0.340 | -1.214 | 1.046 |
| GDP | -0.232 | -0.057 | -0.799 | 1.063 |
| Population density | -0.199 | -0.040 | -0.075 | 0.287 |
| Average smokers | 0.913 | -0.462 | -0.555 | 0.580 |
| Alcohol per capita consumption (15+) in litres | 0.248 | 0.005 | 0.162 | -0.357 |
| ***Environmental Indicators*** |  |  |  |  |
| Air pollution | 0.426 | -0.287 | -0.677 | 0.800 |
| Average annual temperature | -1.011 | 0.567 | 0.717 | -0.829 |
| Water sanitation | 0.139 | -0.113 | -1.181 | 1.251 |
| Air quality | -0.204 | -0.092 | -0.994 | 1.289 |
| Environment pollution index | 0.214 | -0.294 | -1.015 | 1.307 |
| ***Covid-19 Indicators*** |  |  |  |  |
| Skewed distribution | -0.128 | 0.026 | 0.149 | -0.093 |
| Excess death | 0.611 | 0.005 | -0.795 | 0.329 |
| ***Health Indicators*** |  |  |  |  |
| NC Disease Mortality rate (per 100 000) | 0.242 | -0.036 | 0.947 | -1.072 |
| Prevalence of overweight in adults % (BMI >= 25) | 0.230 | 0.451 | -1.282 | 0.416 |
| Probability of dying from Cardio, Cancer, Diabetes, Chronic respiratory (age 30-70) | 0.445 | -0.165 | 0.989 | -1.070 |
| Mortality rate to unsafe WASH | -0.489 | -0.381 | 1.444 | -0.491 |
| Probability of dying from Trachea bronchus, lung cancers (age 30-70) | 1.452 | -0.393 | -0.464 | -0.020 |
| Probability of dying from Lower respiratory infections (age 30-70) | -0.485 | -0.324 | 1.387 | -0.525 |
| Probability of dying from Ischaemic heart disease (age 30-70) | 1.616 | -0.354 | -0.108 | -0.559 |
| Probability of dying from COPD (age 30-70) | 0.398 | -0.376 | 0.556 | -0.277 |

**Table 4.** Clusters of 120 countries using K-means clustering (*k=4*)

| Clusters | Countries |
| --- | --- |
| (a) Cluster 1<br>(28 countries) | Afghanistan, Bangladesh, Botswana, Cambodia, Cameroon, Equatorial Guinea, Eswatini, Ethiopia, Fiji, Ghana, India, Indonesia, Kenya, Madagascar, Malawi, Mozambique, Myanmar, Namibia, Nepal, Nigeria, Pakistan, Philippines, Rwanda, Senegal, Sudan, Uganda, Zambia, Zimbabwe |
| (b) Cluster 2<br>(43 countries) | Algeria, Argentina, Azerbaijan, Bahrain, Belize, Bolivia, Brazil, Colombia, Costa Rica, Dominican Republic, Ecuador, Egypt, El Salvador, Guatemala, Honduras, Iran, Iraq, Jordan, Kazakhstan, Kuwait, Kyrgyzstan, Lebanon, Libya, Malaysia, Maldives, Mexico, Morocco, Oman, Palestine, Panama, Paraguay, Peru, Qatar, Saudi Arabia, Seychelles, South Africa, Sri Lanka, Thailand, Tunisia, Turkey, UAE, Venezuela, Vietnam |
| (c) Cluster 3<br>(21 countries) | Albania, Armenia, Belarus, Bosnia and Herzegovina, Bulgaria, China, Croatia, Cuba, Georgia, Hungary, Latvia, Lithuania, Moldova, Mongolia, North Macedonia, Poland, Romania, Russia, Serbia, Slovenia, Ukraine |
| (d) Cluster 4<br>(28 countries) | Austria, Belgium, Canada, Chile, Cyprus, Czechia, Denmark, Estonia, France, Germany, Greece, Ireland, Israel, Italy, Japan, Luxembourg, Netherlands, Norway, Portugal, South Korea, Singapore, Slovakia, Spain, Sweden, Switzerland, UK, Uruguay, USA |

The second cluster consisted of the 43 countries as in Table 4 (b) and was the third highest in the death-infectivity rate (0.131). Countries like Peru, Brazil, Mexico, Ecuador, are the ones which had a very high death-infectivity rate, however the mean of all the countries in this cluster was close to 2.22%. This cluster was a mixture of countries of different economic status like UAE, Saudi Arabia which were upper-middle economic countries, whereas countries like Iraq, Iran, Bolivia which were low income countries. This cluster has second highest cluster mean of confirmed cases as well as deaths. It can be seen within the cluster that the environmental indicators like air pollution, water sanitation, air quality and EPI have least cluster means. Among health indicators, percentage of overweight in adults is highest among all the clusters. From the Spearman’s correlation in Table 2, it can be observed that, probability of deaths due to different diseases has negative correlation with COVID-19 cases as well as deaths. The minimum cluster means of these features in the second cluster justifies the fact why this cluster ranks second in number of confirmed cases and deaths.

Cluster 3 in this study consists of 21 countries as it can be seen in Table 4 (c). These countries have the highest death-infectivity rate with an average rate of 2.33%. Countries like Slovenia, Bulgaria, Bosnia and Herzegovina, Georgia, Poland, Hungary, are in this clusters and have very high deaths per million people due to COVID-19. However, this cluster is only the third highest in terms of number of cases and last in number of deaths. Further, data shows that these countries are mainly from upper middle and high income countries but with high median age and percentage of senior citizens [48] and that can be one reason why the death-infectivity rate was very high. Other health indicators like probability of deaths due to non-communicable diseases, Cancer, Diabetes, WASH, respiratory infections and COPD had high cluster means.

The last cluster consisted of 28 countries which were all high income countries as shown in Table 4 (d). This cluster topped in both number of confirmed cases as well as deaths, but was last in death-infectivity rate. Countries like USA, UK, Spain, France, Germany, Sweden featured in this clustered. These countries despite of good economy and health care could not sustain to COVID-19 infections and deaths. The positive correlation of COVID-19 cases, deaths and death-infectivity rate with socio-economic factors like median age, percentage of senior citizens, GDP etc. are clearly justified with the high cluster means for these features. The high cluster means with the environmental factors and least cluster means with the health indicators relate to the fact mere having of good environment and health care systems will definitely not protect from COVID-19 effects. The countries which had seen high impact of infections and deaths were those which had high median age and high percentage of senior citizen population. This has indeed appeared in many literatures that elderly people were at highest risk of getting infected [58, 59]. The clusters and the countries within them have also been shown in Figure 6 for better interpretations.

**Figure 6.**
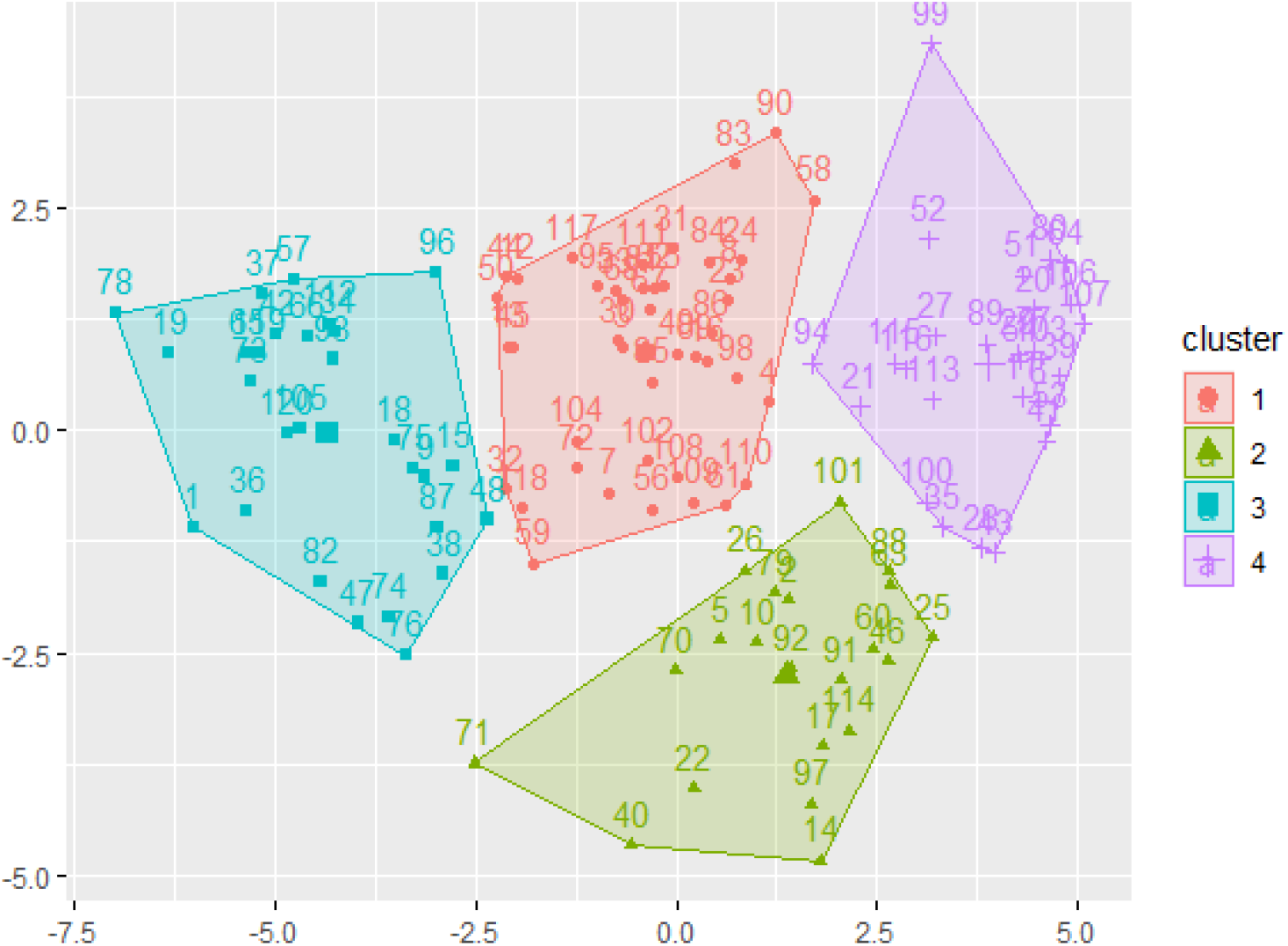
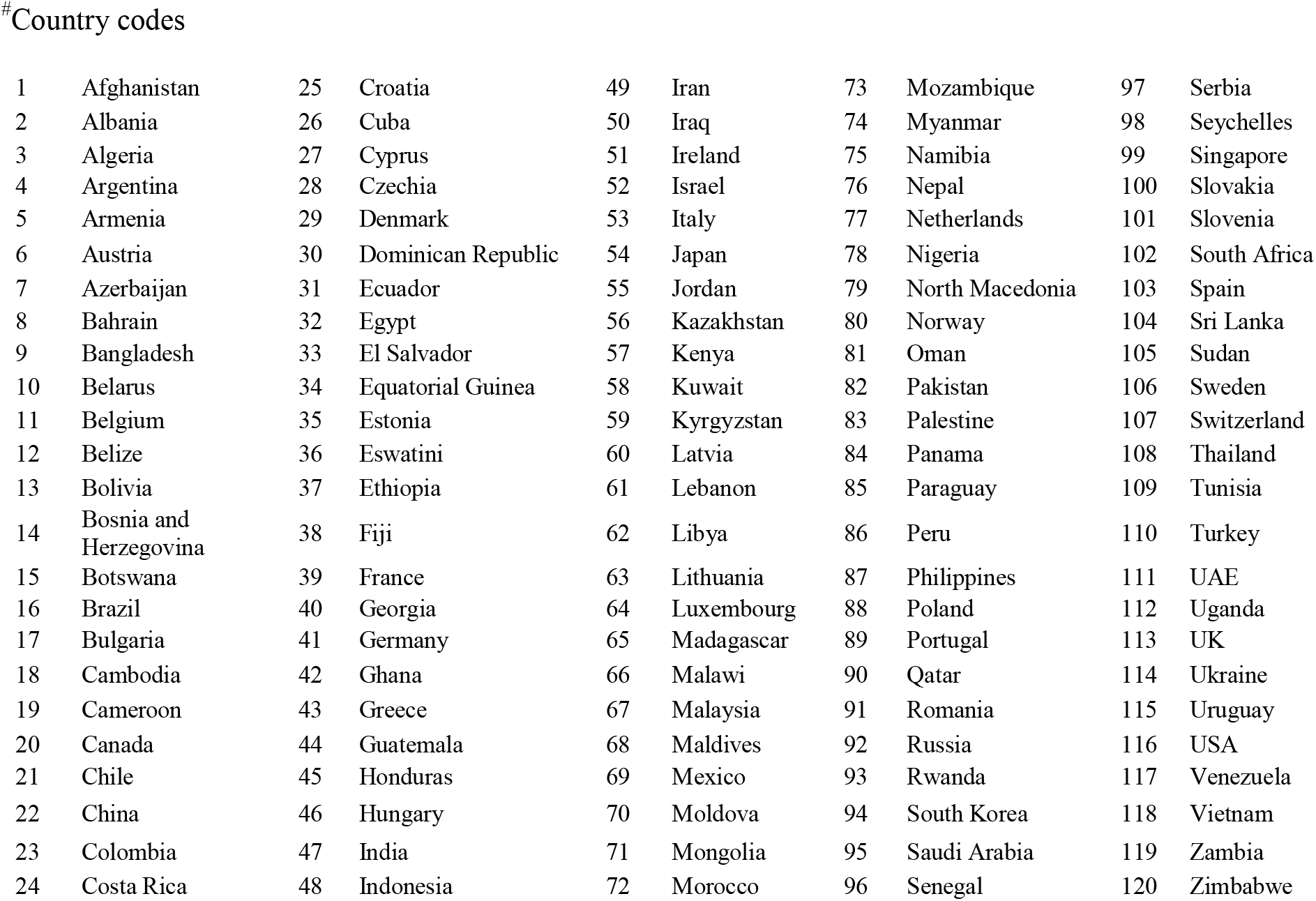
Clusters of most affected 120 countries^#^ considered in this study using K-means clustering

### 3.5 Performance analysis of clustering

In order to analyze the performance of clustering with k-means algorithm in this study, the inertia, Silhouette Coefficient, Calinski-Harabasz index and Davis-Bouldin index were computed. As the ground truth of clusters was unknown in this study, these scores would help to find out how well the clusters were formed with all the parameters taken into consideration.

As three COVID-19 features i.e. death-infectivity rate, number of confirmed cases and number of confirmed deaths were associated with other 23 independent features; performance analysis was done with respect to all the three features. From Table 5, it can be clearly seen that inertia computed from the clustering algorithm were very low in all the three cases where as the Silhouette coefficient index score were very high (≈ 1) in case of number of confirmed cases and The Calinski-Harabasz index scores computed during the analysis were also very high in case of number of confirmed cases as well as deaths; however, comparatively it was low for death-infectivity rate. Finally, very low scores (≈ 0.1) were computed for the Davis-Bouldin index in two cases and moderate score (0.471) for death-infectivity rate. From these scores, it could be clearly depicted that the clusters of 120 countries formed in this study were very well and accurately defined.

**Table 5.** Performance analysis of K-means algorithm in forming the clusters

| Parameters | Death-infectivity rate | Confirmed cases | Confirmed deaths |
| --- | --- | --- | --- |
| Inertia | 0.046 | 0.003 | 0.005 |
| Silhouette Coefficient | 0.606 | 0.971 | 0.979 |
| Calinski-Harabasz Index | 332.29 | 28076.11 | 27292.32 |
| Davies-Bouldin Index | 0.471 | 0.121 | 0.117 |

### 3.6 Correlating clusters with peaks and gaps

With k-means clustering, the 120 countries considered in this study were clustered into 4 groups. It is worth mentioning that the clusters were formed based on the COVID-19 number of confirmed cases, deaths and death-infectivity rates which were associated with 23 features related to socio-economic, environment and health factors. While we correlate these clusters of countries with the waves, peaks and their patterns, it was found that in first cluster, 14 countries have encountered only second wave of the pandemic so far, whereas 6 countries have seen the third wave and Kenya was the only country in the fourth wave. In this cluster, maximum of the countries have experienced 2 peaks so far however, it contained countries Bangladesh and Pakistan which have experienced 5 and 6 peaks respectively. The second cluster of countries have 22 countries which have encountered the second wave so far followed by 6 countries with third wave and 3 countries, Turkey, South Africa and Dominican Republic, have encountered 4 waves of the pandemic so far. This cluster has maximum countries with 3 peaks and 4 peaks. However, there are also countries which have encountered 5 peaks so far. The third cluster consists of 10 countries which have encountered second wave of the pandemic followed by 7 countries experiencing third wave till recent. This cluster had Armenia which has seen four waves of pandemic so far. In this cluster, 11 countries had experienced 3 peaks and 5 countries had 4 peaks of COVID-19 infections. China was the only exceptional case in this country which has reported 1 peak since its inception. The fourth cluster in this study consisted of 14 countries which are in second wave, 6 countries in third wave and Japan with four waves so far. However, there were also 7 countries which have encountered only one wave so far. Among the peaks, maximum of them had encountered 3 and 4 peaks of infections so far. But, there were also countries like USA and Chile, which have 6 peaks and Netherlands with 7 peaks of COVID-19 infections. An overall analysis of all these individual scenarios of the clusters depict that countries which are in cluster 1 and cluster 4 have a probability of seeing highest number of peaks with increase in number of infections, whereas countries in cluster 2 and cluster 3 are expected to have 4 to 5 peaks with increase in the rate of infectivity.

Further, while correlating the waves and peaks with the patterns achieved through skewed distribution, it was observed that for countries which have encountered 2 waves of infections during this period, the intensity of second wave was always higher than the first in most cases. Whereas, for the countries which have encountered three waves so far, the intensity of the third wave was higher among all the three waves. This study found only 6 countries out of 120 which have encountered the fourth wave of pandemic, 3 out of them had highest intensity in second wave and other 3 had high intensity in all other waves. But, if a trend is mapped with the second and third wave experiencing countries, it can be clearly concluded that as the new waves appears, their relative intensities get higher and more peaks can be expected. To justify this fact, with the new variant Omicron showing up, the number of infections is getting higher for almost all the countries where this variant has been found resulting into new COVID-19 waves. Hence, answering to the quest with which the study was carried, it was found that the among the 23 features considered for clustering, the percentage of senior citizens, high median age, unhealthy population, water and sanitation were the factors having higher correlation with COVID-19 cases and death. Hence, these factors are potentials reasons for countries seeing maximum number of infections. Further, with increased number of waves, the consecutive intensities will also definitely increase as compared to other preceding waves.

## 4. Conclusion

In this study, a detailed analysis of COVID-19 features, infectivity (number of confirmed cases), deaths and death-infectivity rate was performed for 120 countries across the world which had highest number of infections and deaths since July 2021. The studies was performed for a period of 23 months (February 2020 – December 2021) to get insights on the association of the infectivity and deaths with socio-economic, environmental and health parameters and identify the potential indicators for possibilities of upcoming COVID-19 peaks. A mathematical model based on mean absolute deviation was built to identify and analyze the peaks and their intensities in all these countries. Alongside, the gaps between the peaks as well as their skewed distribution were computed to identify the rate of growth and decline of the infectivity for each peak. Thereafter, 23 independent features related to socio-economy, environment and health were cross-correlated with COVID-19 features to find the possible association of these factors with the increase and propagation of COVID-19 cases. Further, based on this analysis the countries were clustered into different groups using k-means clustering. These individual clusters helped us to identify the possibility of new peaks which may occur in near future and with their potential indicators and thus answering the question what is going to happen next.

## Data Availability

All data have been taken from open source which are readily available and if needed authors can produce on request.

## Acknowledgement

GNS thanks DST for JC Bose Fellowship.

## References

[1] Dobson, A. P., and E. R. Carper, Infectious diseases and human population history: throughout history the establishment of disease has been a side effect of the growth of civilization. Bioscience, 1996. 46: p. 115–126

[2] Lindahl, J. F. and D. Grace, The consequences of human actions on risks for infectious diseases: a review. Infection ecology & epidemiology, 2015. 5(1): p.30048

[3] Zietz, B. P. and H. Dunkelberg, The history of the plague and the research on the causative agent Yersinia pestis. International journal of hygiene and environmental health, 2004. 207(2): p.165–178

[4] Faruque, S. M., M. J. Albert, and J. J. Mekalanos, Epidemiology, genetics, and ecology of toxigenic Vibrio cholerae. Microbiology and molecular biology reviews, 1998. 62(4): p.1301–1314

[5] Ali, M., A. R. Nelson, A. L. Lopez and D. A. Sack, Updated global burden of cholera in endemic countries. PLoS neglected tropical diseases, 2015. 9(6): p.e0003832.

[6] Cholera Annual Report 2019 Weekly Epidemiological Record 31 2020. 95(38): p. 441–448

[7] Reid, A. H., J. K. Taubenberger and T. G. Fanning, Evidence of an absence: the genetic origins of the 1918 pandemic influenza virus. Nature Reviews Microbiology, 2004. 2(11): p. 909–914

[8] Belser, J. A. and T. M. Tumpey, The 1918 flu, 100 years later. Science, 2018. 359(6373): p. 255–255

[9] Wolfe, N. D., C. P. Dunavan and J. Diamond, Origins of major human infectious diseases. Nature 2007. 447(7142): p.279–283

[10] Piret, J. and G. Boivin, Pandemics throughout history. Frontiers in microbiology, 2021: p. 3594

[11] Huremović, D., Brief history of pandemics (pandemics throughout history). In Psychiatry of pandemics 2019. Springer, Cham, p. 7–35

[12] Caminade, C., K. M. McIntyre and A. E. Jones, Impact of recent and future climate change on vector-borne diseases. Annals of the New York Academy of Sciences, 2019. 1436(1): p.157

[13] Morens, D. M., G. K. Folkers, and A. S. Fauci, The challenge of emerging and re-emerging infectious diseases. Nature, 2004. 430(6996): p. 242–249

[14] Kilpatrick, A. M. and S. E. Randolph, Drivers, dynamics, and control of emerging vector-borne zoonotic diseases. The Lancet, 2012. 380(9857): p.1946–1955

[15] Zhu, N., et al., A novel coronavirus from patients with pneumonia in China, 2019. New England journal of medicine, 2020.

[16] Lau, S. K., et al., Possible bat origin of severe acute respiratory syndrome coronavirus 2. Emerging infectious diseases, 2020. 26(7): p.1542

[17] Lam, T.T.Y., et al., Identifying SARS-CoV-2-related coronaviruses in Malayan pangolins. Nature, 2020. 583(7815): p.282–285

[18] World Health Organization [WHO] (2020e). WHO Coronavirus Disease (COVID-19) Dashboard. Available online at: https://covid19.who.int/ (accessed August 18, 2021).

[19] World Health Organization [WHO] (2021e). WHO Coronavirus Disease (COVID-19) Dashboard. Available online at: https://covid19.who.int/ (accessed December 30, 2021).

[20] Blumenthal, D., E. J. Fowler, M. Abrams and S. R. Collins, COVID-19—implications for the health care system. New England Journal of Medicine, 2020. 383(15): p.1483–1488

[21] Metzl, J. M., A. Maybank and F. De Maio, Responding to the COVID-19 pandemic: the need for a structurally competent health care system. Jama, 2020. 324(3): p.231–232

[22] Khan, J. R., N. Awan, M. Islam and O. Muurlink, Healthcare capacity, health expenditure, and civil society as predictors of COVID-19 case fatalities: a global analysis. Frontiers in public health, 2020. 8: p.347

[23] Gryuk, O., M. Krotovskaya, Y. Borisova, M. Middell, Digital Economy in the Post-COVID Period. 2021. Available at SSRN 3988148.

[24] Khan, N., S. Fahad, M. Naushad and S. Faisal, Analysis of Past and Present Situation of COVID-2019 in the World and its Impact on the World Economy. 2021.Available at SSRN 3858620.

[25] Liu, M., R. Thomadsen and S. Yao, Forecasting the spread of COVID-19 under different reopening strategies. Scientific reports, 2020. 10(1): p.1–8.

[26] Agarwal, P. and K. Jhajharia, Data analysis and modeling of COVID-19. Journal of Statistics and Management Systems, 2021. 24(1): p.1–16.

[27] Pandey, P., et al., A novel fractional mathematical model of COVID-19 epidemic considering quarantine and latent time. Results in Physics, 2021. 26: p.104286.

[28] Nayak, S. R., V. Arora, U. Sinha and R. C. Poonia, A statistical analysis of COVID-19 using Gaussian and probabilistic model. Journal of Interdisciplinary Mathematics, 2021. 24(1): p.19–32.

[29] Diagne, M. L., H. Rwezaura, S. Y. Tchoumi and J. M. Tchuenche, A mathematical model of COVID-19 with vaccination and treatment. Computational and Mathematical Methods in Medicine, 2021.

[30] Sharma, R. K., A. Srivastava, M. Vinay and R. Sethi, An integrated framework of socio-economic and technological interventions for COVID-19 in different economies. Journal of Statistics and Management Systems, 2021. 24(1): p.81–98.

[31] Pal, S. K. and A. K. Pal, The impact of increase in COVID-19 cases with exceptional situation to SDG: Good health and well-being. Journal of Statistics and Management Systems, 2021. 24(1): p.209–228.

[32] Martins-Filho, P. R., Relationship between population density and COVID-19 incidence and mortality estimates: A county-level analysis. Journal of Infection and Public Health, 2021. 14(8): p.1087.

[33] Calina, D., et al., COVID-19 pandemic and alcohol consumption: Impacts and interconnections. Toxicology Reports, 2021

[34] Siddiqui, M. K., et al., Correlation between temperature and COVID-19 (suspected, confirmed and death) cases based on machine learning analysis. J Pure Appl Microbiol, 2020. 14(suppl 1): p.1017–24.

[35] Tsigaris, P. and J. A. Teixeira da Silva, Smoking prevalence and COVID-19 in Europe. Nicotine and Tobacco Research, 2020. 22(9): p.1646–1649.

[36] Wolter, N., et al. Early assessment of the clinical severity of the SARS-CoV-2 Omicron variant in South Africa. medRxiv, 2021

[37] Khan, M. H. R. and A. Hossain, Machine Learning Approaches Reveal That the Number of Tests Do Not Matter to the Prediction of Global Confirmed COVID-19 Cases. Frontiers in artificial intelligence, 2020. 3

[38] Carrillo-Larco, R. M. and M. Castillo-Cara, Using country-level variables to classify countries according to the number of confirmed COVID-19 cases: An unsupervised machine learning approach. Wellcome Open Research, 2020. 5

[39] McCall, B., COVID-19 and artificial intelligence: protecting health-care workers and curbing the spread. The Lancet Digital Health, 2020. 2(4): p.e166–e167

[40] Hu, C., et al., Early prediction of mortality risk among severe COVID-19 patients using machine learning. MedRxiv, 2020.

[41] Ghoshal, B. and A. Tucker, Estimating uncertainty and interpretability in deep learning for coronavirus (COVID-19) detection. arXiv preprint 2003.10769, 2020.

[42] Hu, Z., Ge, Q., Li, S., Jin, L. and Xiong, M., 2020. Artificial intelligence forecasting of COVID-19 in china. arXiv preprint 2002.07112.

[43] Quiroz-Juarez, M.A., Torres-Gomez, A., Hoyo-Ulloa, I., Leon-Montiel, R.D.J. and U’Ren, A.B., 2021. Identification of high-risk COVID-19 patients using machine learning. medRxiv.

[44] Mahdavi, M., et al., A machine learning based exploration of COVID-19 mortality risk. Plos one, 2021. 16(7): p.e0252384.

[45] Chadaga, K., et al., Battling COVID-19 using machine learning: A review. Cogent Engineering, 2021. 8(1): p.1958666.

[46] Worldometer, 2020 COVID-19 Coronavirus. Available at: https://www.worldometers.info/coronavirus/ (Accessed: December, 2021)

[47] Ritchie, H., et al., Coronavirus Pandemic (COVID-19). Published online at http://OurWorldInData.org. 2020, xRetrieved from: ‘ https://ourworldindata.org/coronavirus‘ [Online Resource] (Accessed: December, 2021)

[48] W. Global Health Observatory|, Global health observatory data repository, 2018 URL: https://apps.who.int/gho/data/node.main. (Accessed: August, 2021)

[49] Wendling, Z. A., et.al, Environmental performance index |environmental performance index, 2020, URL: https://epi.yale.edu/epiresults/2020/component/epi (Accessed: August, 2021)

[50] Pham-Gia, T. and T. L. Hung, The mean and median absolute deviations. Mathematical and computer Modelling, 2001. 34(7-8): p.921–936.

[51] Zar, J. H., Spearman rank correlation. Encyclopedia of biostatistics, 2005. 7

[52] Mukaka, M. M., A guide to appropriate use of correlation coefficient in medical research. Malawi medical journal, 2012. 24(3): p.69–71.

[53] MacQueen, J., June. Some methods for classification and analysis of multivariate observations. In Proceedings of the fifth Berkeley symposium on mathematical statistics and probability, 1967. 1(14): p. 281–297

[54] Rizvi, S., M. Umair and M. Cheema, Clustering of countries for COVID-19 cases based on disease prevalence, health systems and environmental indicators. Chaos Solitons Fractals 2021. 151: p-111240.

[55] Dudek, A., 2019, September. Silhouette index as clustering evaluation tool. In Conference of the Section on Classification and Data Analysis of the Polish Statistical Association (pp. 19–33). Springer, Cham.

[56] Shahapure, K. R. and C. Nicholas, Cluster Quality Analysis Using Silhouette Score. In 2020 IEEE 7th International Conference on Data Science and Advanced Analytics (DSAA), 2020. p. 747–748

[57] Liu, Y., et al., Understanding of internal clustering validation measures. In 2010 IEEE international conference on data mining, 2010. p. 911–916

[58] Daoust, J. F., Elderly people and responses to COVID-19 in 27 Countries. PloS one, 2020. 15(7): p.e0235590.

[59] Van Jaarsveld, G. M., The effects of COVID-19 among the elderly population: a case for closing the digital divide. Frontiers in psychiatry, 2020, 11.

