## Supplementary Materials for "COVID-19 impact on Socio-economic and Health Interventions: A Gaps and Peaks analysis using Clustering Approach"

**List of Supplementary tables**

**Supplementary Table 1** Countries grouped on the basis of reaching highest intensity (number of cases) among multiple Covid-19 waves. 25 countries were found to be still in the first wave of Covid-19 and do not appear in this table.

**Supplementary Table 2** Countries grouped on the basis of their skewed distributions of the number of confirmed cases

**Supplementary Table 3** Intensities (No. confirmed of cases) during the peaks for top 120 countries with highest number of cases and deaths from February 2020 – December 2021

**Supplementary Table 4** Gaps (in terms of no. of days) between the peaks for top 120 countries with highest number of cases and deaths from February 2020 – December 2021

**Supplementary Table 5** Duration of each peak (from start to end) in terms of no. of days for top 120 countries with highest number of cases and deaths from February 2020 – December 2021

**Supplementary Table 6** Skewed distribution of each peak for top 120 countries with highest number of cases and deaths from February 2020 – December 2021

**Supplementary Table 1** Countries grouped on the basis of reaching highest intensity (number of cases) among multiple Covid-19 waves. 25 countries were found to be still in the first wave of Covid-19 and do not appear in this table.

| Covid-19 waves | Countries |
|---|---|
| First wave (24 Countries) | USA, France, UK, Spain, Italy, Malaysia, Sweden, Japan, Portugal, Switzerland, Tunisia, UAE, Saudi Arabia, Ecuador, Panama, Azerbaijan, Egypt, Lithuania, Moldova, Qatar, Albania, Belize, Ghana, Malawi, North Macedonia |
| Second Wave (81 Countries) | India, Russia, Turkey, Argentina, Colombia, Iran, Germany, Indonesia, Poland, Mexico, Peru, Netherlands, Czechia, Iraq, Chile, Canada, Belgium, Romania, Hungary, Nepal, Austria, Kazakhstan, Jordan, Morroco, Bolivia, Belarus, Slovakia, Costa Rica, Georgia, Kuwait, Dominican Republic, Guatemala, Palestine, Venezuela, Oman, Ethiopia, Bahrain, Slovenia, Armenia, Libya, Afghanistan, Bosnia and Herzegovina, El Salvador, Estonia, Latvia, Madagascar, Mozambique, Myanmar, Nigeria, Rwanda, Senegal, Singapore, Uganda, Zambia, Zimbabwe |
| Third wave (14 Countries) | Ukraine, Philippines, Bangladesh, Pakistan, Israel, Serbia, Greece, Bulgaria, Croatia, Algeria, Cyprus, Eswatini, Kenya, Kyrgyzstan |
| Fourth wave (1 Country) | South Africa |

**Supplementary Table 2** Countries grouped on the basis of their skewed distributions of the number of confirmed cases

| Skewed distribution | Countries |
|---|---|
| Left distributed (30 Countries) | France, Argentina, Italy, Germany, South Africa, Ukraine, Romania, Japan, Switzerland, Jordan, Panama, Kuwait, Croatia, Denmark, Bahrain, Moldova, Afghanistan, Belize, Botswana, Cambodia, China, El Salvador, Equatorial Guinea, Estonia, Eswatini, Mongolia, Myanmar, Nigeria, North Macedonia, Zimbabwe |
| Right distributed (11 Countries) | Czechia, Nepal, Greece, Slovakia, Sri Lanka, Ethiopia, Armenia, Luxembourg, Maldives, Mozambique, Namibia |
| Mixed distributed (74 Countries) | USA, India, Brazil, Russia, UK, Turkey, Colombia, Spain, Iran, Indonesia, Poland, Mexico, Peru, Netherlands, Iraq, Chile, Philippines, Canada, Bangladesh, Malaysia, Belgium, Sweden, Pakistan, Portugal, Israel, Hungary, Austria, Kazakhstan, Tunisia, Serbia, UAE, Morroco, Saudi Arabia, Lebanon, Ecuador, Bolivia, Paraguay, Belarus, Bulgaria, Costa Rica, Georgia, Thailand, Uruguay, Azerbaijan, Dominican Republic, Guatemala, Palestine, Venezuela, Oman, Egypt, Ireland, Lithuania, Honduras, Cuba, Slovenia, Qatar, Libya, Albania, Algeria, Bosnia and Herzegovina, Cameroon, Ghana, Kenya, Kyrgyzstan, Latvia, Madagascar, Malawi, Norway, South Korea, Singapore, Sudan, Uganda, Vietnam, Zambia |

**Supplementary Table 3** Intensities (No. of confirmed cases) during the peaks for top 120 countries with highest number of deaths and cases from February 2020 – December 2021

| Sl. No. | Country | No. of Peaks | No. of waves | Wave Pattern | Peak 1 (P1) | Peak 2 (P2) | Peak 3 (P3) | Peak 4 (P4) | Peak 5 (P5) | Peak 6 (P6) | Peak 7 (P7) |
|---|---|---|---|---|---|---|---|---|---|---|---|
| 1 | USA | 6 | 2 | A | 48312 | 65073 | 203022 | 303461 | 84337 | 255297 | |
| 2 | India | 4 | 2 | B | 61242 | 81484 | 401993 | 47092 | | | |
| 3 | Brazil | 4 | 1 | | 60713 | 87969 | 88696 | 124248 | | | |
| 4 | Russia | 4 | 2 | B | 17987 | 26613 | 21312 | 39931 | | | |
| 5 | France | 4 | 3 | A | 50746 | 104707 | 117900 | 27781 | | | |
| 6 | UK | 5 | 2 | A | 4534 | 26707 | 68192 | 53980 | 51548 | | |
| 7 | Turkey | 4 | 4 | B | 5138 | 44506 | 63082 | 22332 | | | |
| 8 | Argentina | 3 | 2 | B | 18326 | 13835 | 41080 | | | | |
| 9 | Colombia | 4 | 2 | B | 10284 | 21078 | 19925 | 28315 | | | |
| 10 | Spain | 4 | 2 | A | 9630 | 55019 | 93822 | 61628 | | | |
| 11 | Italy | 3 | 2 | A | 31756 | 26790 | 6613 | | | | |
| 12 | Iran | 3 | 2 | B | 13922 | 25582 | 50228 | | | | |
| 13 | Germany | 2 | 2 | B | 49044 | 31721 | | | | | |
| 14 | Indonesia | 4 | 2 | B | 4823 | 6267 | 14518 | 39532 | | | |
| 15 | Poland | 2 | 3 | B | 21897 | 35253 | | | | | |
| 16 | Mexico | 3 | 2 | B | 9556 | 22339 | 28953 | | | | |
| 17 | South Africa | 3 | 4 | D | 10107 | 21980 | 26485 | | | | |
| 18 | Ukraine | 3 | 3 | C | 15456 | 20456 | 28477 | | | | |
| 19 | Peru | 2 | 2 | B | 21358 | 19026 | | | | | |
| 20 | Netherlands | 7 | 2 | B | 1346 | 11172 | 13072 | 9160 | 9230 | 11297 | 23714 |
| 21 | Czechia | 2 | 2 | B | 15663 | 16816 | | | | | |
| 22 | Iraq | 5 | 2 | B | 4756 | 3701 | 4336 | 8696 | 12734 | | |
| 23 | Chile | 6 | 3 | B | 13188 | 7592 | 4959 | 7592 | 8920 | 2281 | |

| | | | | | | | | | | |
|---|---|---|---|---|---|---|---|---|---|---|
| 24 | Philippines | 3 | 3 | C | 6725 | 10002 | 8537 | | | |
| 25 | Canada | 5 | 3 | B | 2567 | 7828 | 10643 | 10985 | 11381 | |
| 26 | Bangladesh | 5 | 3 | C | 4014 | 3200 | 2316 | 7626 | 15989 | |
| 27 | Malaysia | 3 | 1 | A | 5728 | 9020 | 21176 | | | |
| 28 | Belgium | 4 | 2 | B | 23921 | 9065 | 5010 | 47836 | | |
| 29 | Sweden | 3 | 1 | A | 32485 | 21802 | 1355 | | | |
| 30 | Romania | 3 | 2 | B | 10269 | 6115 | 18863 | | | |
| 31 | Pakistan | 6 | 3 | C | 12073 | 3306 | 4974 | 8495 | 5026 | 7727 |
| 32 | Japan | 3 | 4 | A | 7957 | 7238 | 20028 | | | |
| 33 | Portugal | 4 | 2 | A | 1516 | 8371 | 16432 | 4376 | | |
| 34 | Israel | 3 | 3 | C | 9078 | 11934 | 22291 | | | |
| 35 | Hungary | 3 | 3 | B | 6697 | 11265 | 27830 | | | |
| 36 | Switzerland | 4 | 2 | A | 1163 | 21926 | 5583 | 12801 | | |
| 37 | Nepal | 3 | 2 | B | 5743 | 5657 | 3194 | | | |
| 38 | Austria | 3 | 2 | B | 9586 | 3895 | 15809 | | | |
| 39 | Kazakhstan | 3 | 2 | B | 18757 | 3856 | 66121 | | | |
| 40 | Tunisia | 4 | 1 | A | 5752 | 4170 | 1847 | 9823 | | |
| 41 | Jordan | 2 | 3 | B | 7933 | 9535 | | | | |
| 42 | Serbia | 4 | 3 | C | 7780 | 9983 | 8467 | 7111 | | |
| 43 | UAE | 4 | 1 | A | 1578 | 3966 | 4471 | 985 | | |
| 44 | Morroco | 3 | 2 | B | 6195 | 740 | 12039 | | | |
| 45 | Saudi Arabia | 4 | 2 | A | 4919 | 473 | 270 | 5439 | | |
| 46 | Lebanon | 3 | 1 | | 6154 | 3562 | 2591 | | | |
| 47 | Ecuador | 5 | 2 | A | 11536 | 2249 | 3942 | 5913 | 4953 | |
| 48 | Bolivia | 3 | 2 | B | 2031 | 2866 | 7072 | | | |
| 49 | Greece | 3 | 3 | C | 3316 | 4322 | 4327 | | | |
| 50 | Paraguay | 3 | 1 | | 1162 | 3836 | 3481 | | | |
| 51 | Belarus | 4 | 2 | B | 1691 | 1799 | 1756 | 1862 | | |

| | | | | | | | | | |
|---|---|---|---|---|---|---|---|---|---|
| 52 | Bulgaria | 3 | 3 | C | 4828 | 5176 | 6816 | | |
| 53 | Panama | 3 | 2 | A | 1420 | 5186 | 1249 | | |
| 54 | Slovakia | 2 | 2 | B | 11915 | 6103 | | | |
| 55 | Costa Rica | 4 | 2 | B | 1907 | 2772 | 5992 | 6952 | |
| 56 | Georgia | 4 | 2 | B | 5450 | 1792 | 6208 | 5739 | |
| 57 | Kuwait | 4 | 2 | B | 900 | 903 | 1716 | 1993 | |
| 58 | Thailand | 4 | 1 | | 9635 | 18912 | 20533 | 16130 | |
| 59 | Uruguay | 2 | 1 | | 4213 | 315 | | | |
| 60 | Croatia | 3 | 3 | C | 4620 | 3217 | 7315 | | |
| 61 | Azerbaijan | 3 | 3 | A | 4451 | 2237 | 7319 | | |
| 62 | Dominican Republic | 3 | 4 | B | 2370 | 1765 | 1248 | | |
| 63 | Guatemala | 3 | 2 | B | 1739 | 2013 | 5826 | | |
| 64 | Palestine | 5 | 3 | B | 2516 | 1703 | 2884 | 2466 | 30356 |
| 65 | Denmark | 3 | 1 | | 4508 | 2007 | 1036 | | |
| 66 | Venezuela | 4 | 2 | B | 1008 | 673 | 1348 | 2850 | |
| 67 | Oman | 4 | 2 | B | 2164 | 2685 | 3544 | 5517 | |
| 68 | Egypt | 5 | 3 | A | 1503 | 1418 | 1021 | 956 | 741 |
| 69 | Ireland | 3 | 1 | | 1284 | 8227 | 1501 | | |
| 70 | Sri Lanka | 2 | 1 | | 3623 | 9912 | | | |
| 71 | Lithuania | 3 | 2 | A | 3984 | 1522 | 3622 | | |
| 72 | Honduras | 4 | 1 | | 1087 | 1386 | 2720 | 4043 | |
| 73 | Ethiopia | 2 | 2 | B | 1829 | 2173 | | | |
| 74 | Bahrain | 2 | 2 | B | 3273 | 139 | | | |
| 75 | Cuba | 3 | 1 | | 1051 | 9323 | 5049 | | |
| 76 | Slovenia | 3 | 2 | B | 3354 | 1526 | 4521 | | |
| 77 | Moldova | 3 | 2 | A | 1766 | 2273 | 2068 | | |
| 78 | Armenia | 2 | 4 | B | 2474 | 1257 | | | |

| | | | | | | | | |
|---|---|---|---|---|---|---|---|---|
| 79 | Qatar | 4 | 2 | A | 1901 | 273 | 650 | 306 |
| 80 | Libya | 3 | 2 | B | 1639 | 1350 | 4061 | |
| 81 | Afghanistan | 2 | 2 | B | 377 | 1940 | | |
| 82 | Albania | 3 | 2 | A | 892 | 1054 | 1131 | |
| 83 | Algeria | 4 | 3 | C | 953 | 459 | 286 | 1927 |
| 84 | Belize | 2 | 2 | A | 25 | 815 | | |
| 85 | Bosnia | 4 | 3 | B | 1921 | 3755 | 1722 | 2512 |
| 86 | Botswana | 2 | 1 | | 2356 | 8530 | | |
| 87 | Cambodia | 1 | 1 | | 978 | | | |
| 88 | Cameroon | 3 | 1 | | 1183 | 632 | 9668 | |
| 89 | China | 1 | 1 | | 15133 | | | |
| 90 | Cyprus | 0 | 3 | C | | | | |
| 91 | El Salvador | 1 | 2 | B | 2324 | | | |
| 92 | Equatorial Guinea | 2 | 1 | | 1750 | 742 | | |
| 93 | Estonia | 2 | 2 | B | 1956 | 2300 | | |
| 94 | Eswatini | 1 | 3 | C | 1068 | | | |
| 95 | Fiji | 0 | 1 | | | | | |
| 96 | Ghana | 3 | 3 | A | 1385 | 655 | 1583 | |
| 97 | Kenya | 3 | 4 | C | 1554 | 2008 | 1974 | |
| 98 | Kyrgyzstan | 3 | 3 | C | 1030 | 1965 | 954 | |
| 99 | Latvia | 3 | 2 | B | 1861 | 1036 | 3206 | |
| 100 | Luxembourg | 3 | 1 | | 1967 | 608 | 398 | |
| 101 | Madagascar | 2 | 2 | B | 859 | 712 | | |
| 102 | Malawi | 1 | 2 | A | 1316 | | | |
| 103 | Maldives | 1 | 1 | | 2194 | | | |
| 104 | Mongolia | 2 | 1 | | 2527 | 7394 | | |
| 105 | Mozambique | 1 | 2 | B | 1611 | | | |
| 106 | Myanmar | 2 | 2 | B | 1527 | 7083 | | |

| | | | | | | | | | |
|---|---|---|---|---|---|---|---|---|---|
| 107 | Namibia | 1 | 1 | | 3268 | | | | |
| 108 | Nigeria | 2 | 3 | B | 1634 | 1823 | | | |
| 109 | North Macedonia | 4 | 2 | A | 1402 | 1511 | 1250 | 677 | |
| 110 | Norway | 4 | 1 | | 1680 | 935 | 1150 | 1785 | |
| 111 | Rwanda | 1 | 2 | B | 595 | | | | |
| 112 | S.Korea | 4 | 1 | | 1237 | 797 | 1896 | 3270 | |
| 113 | Senegal | 0 | 2 | B | | | | | |
| 114 | Seychelles | 0 | 1 | | | | | | |
| 115 | Singapore | 5 | 2 | B | 1426 | 908 | 58 | 45 | 2478 |
| 116 | Sudan | 2 | 2 | A | 1099 | 390 | | | |
| 117 | Uganda | 2 | 2 | B | 1859 | 1735 | | | |
| 118 | Vietnam | 2 | 1 | | 17428 | 39132 | | | |
| 119 | Zambia | 2 | 2 | B | 1796 | 3594 | | | |
| 120 | Zimbabwe | 2 | 2 | B | 1365 | 3110 | | | |

**Supplementary Table 4** Gaps (in terms of no. of days) between the peaks for top 120 countries with highest number of deaths and cases from February 2020 – December 2021

| Sl. No. | Country | No. of peaks | $\Delta G_1 = T_{P2} - T_{P1}$ | $\Delta G_2 = T_{P3} - T_{P2}$ | $\Delta G_3 = T_{P4} - T_{P3}$ | $\Delta G_4 = T_{P5} - T_{P4}$ | $\Delta G_3 = T_{P4} - T_{P3}$ | $\Delta G_4 = T_{P5} - T_{P4}$ |
|---|---|---|---|---|---|---|---|---|
| 1 | USA | 6 | 184 | 62 | 89 | 89 | 123 | |
| 2 | India | 4 | 92 | 181 | 153 | | | |
| 3 | Brazil | 4 | 153 | 89 | 61 | | | |
| 4 | Russia | 4 | 92 | 150 | 123 | | | |
| 5 | France | 4 | 214 | 151 | 123 | | | |
| 6 | UK | 5 | 153 | 92 | 181 | 91 | | |
| 7 | Turkey | 4 | 153 | 251 | 92 | | | |
| 8 | Argentina | 3 | 92 | 120 | | | | |
| 9 | Colombia | 4 | 215 | 89 | 122 | | | |
| 10 | Spain | 4 | 213 | 61 | 183 | | | |
| 11 | Italy | 3 | 151 | 61 | | | | |
| 12 | Iran | 3 | 120 | 245 | | | | |
| 13 | Germany | 2 | 120 | | | | | |
| 14 | Indonesia | 4 | 61 | 62 | 212 | | | |
| 15 | Poland | 2 | 181 | | | | | |
| 16 | Mexico | 3 | 150 | 212 | | | | |
| 17 | South Africa | 3 | 153 | 181 | | | | |
| 18 | Ukraine | 3 | 120 | 214 | | | | |
| 19 | Peru | 2 | 212 | | | | | |
| 20 | Netherlands | 7 | 184 | 61 | 90 | 61 | 61 | 122 |
| 21 | Czechia | 2 | 151 | | | | | |
| 22 | Iraq | 5 | 61 | 89 | 61 | 123 | | |
| 23 | Chile | 6 | 122 | 123 | 59 | 91 | 122 | |

| | | | | | | | |
|---|---|---|---|---|---|---|---|
| 24 | Philippines | 3 | 212 | 214 | | | |
| 25 | Canada | 5 | 214 | 61 | 120 | 122 | |
| 26 | Bangladesh | 5 | 61 | 122 | 120 | 123 | |
| 27 | Malaysia | 3 | 120 | 61 | | | |
| 28 | Belgium | 4 | 151 | 153 | 91 | | |
| 29 | Sweden | 3 | 120 | 121 | | | |
| 30 | Romania | 3 | 151 | 184 | | | |
| 31 | Pakistan | 6 | 153 | 121 | 61 | 61 | 61 |
| 32 | Japan | 3 | 120 | 122 | | | |
| 33 | Portugal | 4 | 214 | 61 | 181 | | |
| 34 | Israel | 3 | 123 | 222 | | | |
| 35 | Hungary | 3 | 90 | 244 | | | |
| 36 | Switzerland | 4 | 214 | 151 | 153 | | |
| 37 | Nepal | 3 | 181 | 123 | | | |
| 38 | Austria | 3 | 121 | 244 | | | |
| 39 | Kazakhstan | 3 | 304 | 62 | | | |
| 40 | Tunisia | 4 | 92 | 59 | 122 | | |
| 41 | Jordan | 2 | 121 | | | | |
| 42 | Serbia | 4 | 121 | 183 | | | |
| 43 | UAE | 4 | 92 | 150 | 92 | | |
| 44 | Morroco | 3 | 151 | 123 | | | |
| 45 | Saudi Arabia | 4 | 153 | 90 | 112 | | |
| 46 | Lebanon | 3 | 89 | 123 | | | |
| 47 | Ecuador | 5 | 153 | 123 | 89 | 71 | |
| 48 | Bolivia | 3 | 153 | 150 | | | |
| 49 | Greece | 3 | 121 | 122 | | | |
| 50 | Paraguay | 3 | 89 | 61 | | | |

| | | | | | | |
|---|---|---|---|---|---|---|
| 51 | Belarus | 4 | 90 | 92 | 92 | |
| 52 | Bulgaria | 3 | 121 | 183 | | |
| 53 | Panama | 3 | 153 | 150 | | |
| 54 | Slovakia | 2 | 90 | | | |
| 55 | Costa Rica | 4 | 92 | 151 | 122 | |
| 56 | Georgia | 4 | 120 | 123 | 61 | |
| 57 | Kuwait | 4 | 61 | 121 | 122 | |
| 58 | Thailand | 4 | 61 | 60 | | |
| 59 | Uruguay | 2 | 123 | | | |
| 60 | Croatia | 3 | 120 | 214 | | |
| 61 | Azerbaijan | 3 | 90 | 183 | | |
| 62 | Dominican Republic | 3 | 120 | 169 | | |
| 63 | Guatemala | 3 | 89 | 123 | | |
| 64 | Palestine | 5 | 60 | 61 | 122 | 62 |
| 65 | Denmark | 3 | 90 | 91 | | |
| 66 | Venezuela | 4 | 92 | 59 | 183 | |
| 67 | Oman | 4 | 92 | 181 | 61 | |
| 68 | Egypt | 5 | 153 | 120 | 61 | |
| 69 | Ireland | 3 | 92 | 181 | | |
| 70 | Sri Lanka | 2 | 92 | | | |
| 71 | Lithuania | 3 | 151 | 153 | | |
| 72 | Honduras | 4 | 123 | 89 | 123 | |
| 73 | Ethiopia | 2 | 212 | | | |
| 74 | Bahrain | 2 | 122 | | | |
| 75 | Cuba | 3 | 122 | 92 | | |
| 76 | Slovenia | 3 | 89 | 214 | | |
| 77 | Moldova | 3 | 90 | 214 | | |

| | | | | | |
|---|---|---|---|---|---|
| 78 | Armenia | 2 | 151 | | |
| 79 | Qatar | 4 | 123 | 212 | 92 |
| 80 | Libya | 3 | 181 | 92 | |
| 81 | Afghanistan | 2 | 243 | | |
| 82 | Albania | 3 | 153 | | |
| 83 | Algeria | 4 | 59 | 61 | 92 |
| 84 | Belize | 2 | 184 | | |
| 85 | Bosnia and Herzegovina | 4 | 120 | 184 | 61 |
| 86 | Botswana | 2 | 153 | | |
| 87 | Cambodia | 1 | | | |
| 88 | Cameroon | 3 | 152 | 152 | |
| 89 | China | 1 | | | |
| 90 | Cyprus | 0 | | | |
| 91 | El Salvador | 1 | | | |
| 92 | Equatorial Guinea | 2 | 304 | | |
| 93 | Estonia | 2 | 244 | | |
| 94 | Eswatini | 1 | | | |
| 95 | Fiji | 0 | | | |
| 96 | Ghana | 3 | 122 | 62 | |
| 97 | Kenya | 3 | 121 | 153 | |
| 98 | Kyrgyzstan | 3 | 122 | 182 | 61 |
| 99 | Latvia | 3 | 150 | 153 | |
| 100 | Luxembourg | 3 | 121 | 122 | |
| 101 | Madagascar | 2 | 184 | | |
| 102 | Malawi | 1 | 181 | | |
| 103 | Maldives | 1 | | | |
| 104 | Mongolia | 2 | 253 | | |

| | | | | | | |
|---|---|---|---|---|---|---|
| 105 | Mozambique | 1 | | | | |
| 106 | Myanmar | 2 | 212 | | | |
| 107 | Namibia | 1 | -21 | | | |
| 108 | Nigeria | 2 | 183 | | | |
| 109 | North Macedonia | 4 | 120 | 153 | 91 | |
| 110 | Norway | 4 | 62 | 59 | 153 | |
| 111 | Rwanda | 1 | | | | |
| 112 | South Korea | 4 | 89 | 92 | 61 | |
| 113 | Senegal | 0 | | | | |
| 114 | Seychelles | 0 | | | | |
| 115 | Singapore | 5 | 123 | 153 | 89 | 91 |
| 116 | Sudan | 2 | 120 | -203 | | |
| 117 | Uganda | 2 | 181 | | | |
| 118 | Vietnam | 2 | 91 | | | |
| 119 | Zambia | 2 | 150 | | | |
| 120 | Zimbabwe | 2 | 181 | | | |

**Supplementary Table 5** Duration of each peak (from start to end) in terms of no. of days for top 120 countries with highest number of deaths and cases from February 2020 – December 2021

| Sl. No. | Country | No. of peaks | $\delta_1 = L_2 - L_1$ | $\delta_2 = L_3 - L_2$ | $\delta_3 = L_4 - L_3$ | $\delta_4 = L_5 - L_4$ | $\delta_5 = L_6 - L_5$ | $\delta_6 = L_7 - L_6$ | $\delta_6 = L_8 - L_7$ |
|---|---|---|---|---|---|---|---|---|---|
| 1 | USA | 6 | 183 | 92 | 90 | 122 | 123 | 61 | |
| 2 | India | 4 | 183 | 151 | 123 | 153 | | | |
| 3 | Brazil | 4 | 214 | 120 | 92 | 245 | | | |
| 4 | Russia | 4 | 245 | 120 | 123 | 153 | | | |
| 5 | France | 4 | 61 | 214 | 181 | 184 | | | |
| 6 | UK | 5 | 122 | 122 | 182 | 123 | 122 | | |
| 7 | Turkey | 4 | 122 | 212 | 122 | 185 | | | |
| 8 | Argentina | 3 | 184 | 59 | 306 | | | | |
| 9 | Colombia | 4 | 122 | 182 | 91 | 184 | | | |
| 10 | Spain | 4 | 275 | 184 | 181 | 123 | | | |
| 11 | Italy | 3 | 214 | 151 | 122 | | | | |
| 12 | Iran | 3 | 306 | 150 | 215 | | | | |
| 13 | Germany | 2 | 334 | 94 | | | | | |
| 14 | Indonesia | 4 | 123 | 61 | 151 | 214 | | | |
| 15 | Poland | 2 | 123 | 212 | | | | | |
| 16 | Mexico | 3 | 153 | 274 | 153 | | | | |
| 17 | South Africa | 3 | 153 | 181 | 215 | | | | |
| 18 | Ukraine | 3 | 215 | 212 | 122 | | | | |
| 19 | Peru | 2 | 214 | 274 | 92 | | | | |
| 20 | Netherlands | 7 | 153 | 91 | 90 | 61 | 61 | 62 | 122 |
| 21 | Czechia | 2 | 151 | 154 | | | | | |
| 22 | Iraq | 5 | 92 | 121 | 59 | 92 | 153 | | |
| 23 | Chile | 6 | 123 | 91 | 90 | 61 | 153 | 92 | |

| | | | | | | | | |
|---|---|---|---|---|---|---|---|---|
| 24 | Philippines | 3 | 214 | 243 | 153 | | | |
| 25 | Canada | 5 | 91 | 184 | 59 | 184 | 96 | |
| 26 | Bangladesh | 5 | 61 | 92 | 119 | 123 | 214 | |
| 27 | Malaysia | 3 | 151 | 91 | 62 | 91 | | |
| 28 | Belgium | 4 | 334 | 153 | 92 | 92 | | |
| 29 | Sweden | 3 | 243 | 152 | 152 | | | |
| 30 | Romania | 3 | 212 | 123 | 153 | | | |
| 31 | Pakistan | 6 | 153 | 151 | 61 | 61 | 62 | 122 |
| 32 | Japan | 3 | 243 | 122 | 153 | | | |
| 33 | Portugal | 4 | 91 | 184 | 151 | 92 | 92 | |
| 34 | Israel | 3 | 275 | 213 | 122 | | | |
| 35 | Hungary | 3 | 123 | 151 | 153 | | | |
| 36 | Switzerland | 4 | 91 | 243 | 153 | 91 | | |
| 37 | Nepal | 3 | 181 | 152 | 153 | | | |
| 38 | Austria | 3 | 365 | 123 | 153 | | | |
| 39 | Kazakhstan | 3 | 184 | 181 | 62 | | | |
| 40 | Tunisia | 4 | 92 | 59 | 92 | 92 | | |
| 41 | Jordan | 2 | 123 | 181 | | | | |
| 42 | Serbia | 4 | 92 | 150 | 123 | | | |
| 43 | UAE | 4 | 61 | 151 | 123 | 123 | | |
| 44 | Morroco | 3 | 243 | 92 | 153 | | | |
| 45 | Saudi Arabia | 4 | 184 | 61 | 90 | 122 | | |
| 46 | Lebanon | 3 | 151 | 153 | 122 | | | |
| 47 | Ecuador | 5 | 113 | 122 | 90 | 61 | 132 | |
| 48 | Bolivia | 3 | 153 | 121 | 153 | | | |
| 49 | Greece | 3 | 92 | 150 | 62 | | | |
| 50 | Paraguay | 3 | 151 | 92 | 92 | | | |
| 51 | Belarus | 4 | 61 | 90 | 122 | 122 | | |

| | | | | | | | |
|---|---|---|---|---|---|---|---|
| 52 | Bulgaria | 3 | 92 | 150 | 151 | | |
| 53 | Panama | 3 | 92 | 123 | 212 | | |
| 54 | Slovakia | 2 | 151 | 122 | | | |
| 55 | Costa Rica | 4 | 61 | 120 | 153 | 153 | |
| 56 | Georgia | 4 | 151 | 92 | 92 | 92 | |
| 57 | Kuwait | 4 | 153 | 61 | 120 | 123 | |
| 58 | Thailand | 4 | 150 | 123 | 61 | 61 | |
| 59 | Uruguay | 2 | 181 | 91 | | | |
| 60 | Croatia | 3 | 120 | 184 | 153 | | |
| 61 | Azerbaijan | 3 | 120 | 153 | 184 | | |
| 62 | Dominican Republic | 3 | 243 | 153 | 122 | | |
| 63 | Guatemala | 3 | 212 | 123 | 214 | | |
| 64 | Palestine | 5 | 62 | 59 | 153 | 60 | 61 |
| 65 | Denmark | 3 | 120 | 61 | 123 | | |
| 66 | Venezuela | 4 | 91 | 90 | 61 | 244 | |
| 67 | Oman | 4 | 92 | 122 | 151 | 92 | |
| 68 | Egypt | 5 | 122 | 183 | 61 | 61 | 153 |
| 69 | Ireland | 3 | 214 | 182 | 92 | | |
| 70 | Sri Lanka | 2 | 92 | 122 | | | |
| 71 | Lithuania | 3 | 151 | 153 | 153 | | |
| 72 | Honduras | 4 | 123 | 151 | 92 | 214 | |
| 73 | Ethiopia | 2 | 141 | 212 | | | |
| 74 | Bahrain | 2 | 122 | 92 | | | |
| 75 | Cuba | 3 | 90 | 123 | 92 | | |
| 76 | Slovenia | 3 | 151 | 153 | 153 | | |
| 77 | Moldova | 3 | 123 | 181 | 184 | | |
| 78 | Armenia | 2 | 138 | 184 | | | |
| 79 | Qatar | 4 | 122 | 92 | 212 | 153 | |

| | | | | | | |
|---|---|---|---|---|---|---|
| 80 | Libya | 3 | 92 | 181 | 62 | |
| 81 | Afghanistan | 2 | 212 | 306 | | |
| 82 | Albania | 3 | 120 | 123 | | |
| 83 | Algeria | 4 | 62 | 59 | 91 | 62 |
| 84 | Belize | 2 | 141 | 115 | | |
| 85 | Bosnia and Herzegovina | 4 | 61 | 153 | 92 | 61 |
| 86 | Botswana | 2 | 181 | 184 | | |
| 87 | Cambodia | 1 | 62 | | | |
| 88 | Cameroon | 3 | 139 | 61 | 212 | |
| 89 | China | 1 | 34 | | | |
| 90 | Cyprus | 0 | | | | |
| 91 | El Salvador | 1 | 214 | | | |
| 92 | Equatorial Guinea | 2 | 122 | 304 | | |
| 93 | Estonia | 2 | 181 | 184 | | |
| 94 | Eswatini | 1 | 47 | | | |
| 95 | Fiji | 0 | | | | |
| 96 | Ghana | 3 | 123 | 61 | 182 | |
| 97 | Kenya | 3 | 92 | 90 | 183 | |
| 98 | Kyrgyzstan | 3 | 212 | 153 | 153 | |
| 99 | Latvia | 3 | 106 | 153 | 153 | |
| 100 | Luxembourg | 3 | 92 | 150 | 62 | |
| 101 | Madagascar | 2 | 184 | 61 | | |
| 102 | Malawi | 1 | 137 | 87 | | |
| 103 | Maldives | 1 | 114 | | | |
| 104 | Mongolia | 2 | 58 | 92 | | |
| 105 | Mozambique | 1 | 132 | | | |
| 106 | Myanmar | 2 | 151 | 153 | | |
| 107 | Namibia | 1 | | | | |

| | | | | | | | |
|---|---|---|---|---|---|---|---|
| 108 | Nigeria | 2 | 120 | 183 | | | |
| 109 | North Macedonia | 4 | 120 | 153 | 106 | 51 | |
| 110 | Norway | 4 | 45 | 59 | 153 | 92 | |
| 111 | Rwanda | 1 | 91 | | | | |
| 112 | South Korea | 4 | 31 | 89 | 123 | 61 | |
| 113 | Senegal | 0 | | | | | |
| 114 | Seychelles | 0 | | | | | |
| 115 | Singapore | 5 | 102 | 93 | 151 | 91 | 62 |
| 116 | Sudan | 2 | 22 | 53 | -234 | | |
| 117 | Uganda | 2 | 23 | 243 | | | |
| 118 | Vietnam | 2 | 92 | 61 | | | |
| 119 | Zambia | 2 | 24 | 212 | | | |
| 120 | Zimbabwe | 2 | 86 | 244 | | | |

**Supplementary Table 6** Skewed distribution of each peak for top 120 countries with highest number of cases and deaths from February 2020 – December 2021

| Sl. No. | Country | No. of waves | No. of Peaks | Peaks | $\sigma_L$ | $\sigma_R$ | $T_L$ | $T_R$ | $AUC_L$ | $AUC_R$ | $\gamma$ | Peak Distribution | Pattern |
|---|---|---|---|---|---|---|---|---|---|---|---|---|---|
| 1 | USA | 2 | 6 | P1 | 2639255 | 4593265 | 0.10784314 | 0.89215686 | 284625.54 | 4097912.9 | 0.06945622 | **R** | Mixed |
| | | | | P2 | 6435200 | 6429546 | 0.34408602 | 0.65591398 | 2214262.4 | 4217229.1 | 0.52505148 | **R** | |
| | | | | P3 | 6149098 | 4215774 | 0.60927152 | 0.39072848 | 3746470.3 | 1647223 | 2.27441604 | **L** | |
| | | | | P4 | 1885745 | 1314109 | 0.5 | 0.5 | 942872.5 | 657054.5 | 1.43499892 | **L** | |
| | | | | P5 | 1891302 | 1321474 | 0.22764228 | 0.77235772 | 430540.29 | 1020650.7 | 0.42182924 | **R** | |
| | | | | P6 | 5608427 | 6655228 | 0.45901639 | 0.54098361 | 2574359.9 | 3600369.2 | 0.71502664 | **R** | |
| 2 | India | 2 | 4 | P1 | 1661125 | 4616596 | 0.502732 | 0.497268 | 835100.69 | 2295685.5 | 0.36376965 | **R** | Mixed |
| | | | | P2 | 1871498 | 2928159 | 0.006623 | 0.993377 | 12394.931 | 2908765.8 | 0.00426123 | **R** | |
| | | | | P3 | 8052728 | 13645876 | 0.16847826 | 0.83152174 | 1356709.6 | 11346843 | 0.11956715 | **R** | |
| | | | | P4 | 2110883 | 1055333 | 0.39869281 | 0.60130719 | 841593.88 | 634579.32 | 1.32622329 | **L** | |
| 3 | Brazil | 1 | 4 | P1 | 3902555 | 1627333 | 0.71495327 | 0.28504673 | 2790144.5 | 463865.95 | 6.0149801 | **L** | Mixed |
| | | | | P2 | 3669126 | 1346528 | 0.76666667 | 0.23333333 | 2812996.6 | 314189.86 | 8.95317434 | **L** | |
| | | | | P3 | 4107752 | 1886543 | 0.66304348 | 0.33695652 | 2723618.2 | 635682.96 | 4.28455431 | **L** | |
| | | | | P4 | 2011587 | 3078744 | 0.24390244 | 0.75609756 | 490630.98 | 2327830.8 | 0.21076745 | **R** | |
| 4 | Russia | 2 | 4 | P1 | 1499769 | 1521080 | 0.75102041 | 0.24897959 | 1126357.1 | 378717.87 | 2.97413247 | **L** | Mixed |
| | | | | P2 | 681001 | 942407 | 0.25833333 | 0.74166667 | 175925.26 | 698951.86 | 0.25169867 | **R** | |
| | | | | P3 | 436082 | 735655 | 0.49593496 | 0.50406504 | 216268.31 | 370817.97 | 0.58321961 | **R** | |
| | | | | P4 | 2192735 | 1090656 | 0.50204082 | 0.49795918 | 1100842.5 | 543102.17 | 2.0269528 | **L** | |
| 5 | France | 3 | 4 | P1 | 116583 | 22114 | 0.49180328 | 0.50819672 | 57335.902 | 11238.262 | 5.10184764 | **L** | Left |
| | | | | P2 | 13269 | 22995 | 0.98360656 | 0.01639344 | 13051.475 | 376.96715 | 34.6223148 | **L** | |
| | | | | P3 | 2049635 | 400792 | 0.99346405 | 0.00653595 | 2036238.7 | 2619.5565 | 777.321928 | **L** | |
| | | | | P4 | 3000169 | 508097 | 0.67391304 | 0.32608696 | 2021853 | 165683.8 | 12.2030818 | **L** | |

| | | | | | | | | | | | | | |
|---|---|---|---|---|---|---|---|---|---|---|---|---|---|
| 6 | UK | 2 | 5 | P1 | 218724 | 47254 | 0.5 | 0.5 | 109362 | 23627 | 4.62868752 | **L** | Mixed |
| | | | | P2 | 710002 | 618941 | 0.99180328 | 0.00819672 | 704182.31 | 5073.2861 | 138.802012 | **L** | |
| | | | | P3 | 2194451 | 675044 | 0.51098901 | 0.48901099 | 1121340.3 | 330103.93 | 3.39693117 | **L** | |
| | | | | P4 | 1380190 | 937935 | 0.49593496 | 0.50406504 | 684484.47 | 472780.24 | 1.44778569 | **L** | |
| | | | | P5 | 2270510 | 1176255 | 0.23770492 | 0.76229508 | 539711.39 | 896653.4 | 0.60191752 | **R** | |
| 7 | Turkey | 4 | 4 | P1 | 106673 | 110669 | 0.24590164 | 0.75409836 | 26231.066 | 83455.311 | 0.31431272 | **R** | Mixed |
| | | | | P2 | 407974 | 1239516 | 0.5754717 | 0.4245283 | 234777.49 | 526209.62 | 0.44616724 | **R** | |
| | | | | P3 | 2119003 | 605061 | 0.5 | 0.5 | 1059501.5 | 302530.5 | 3.50213119 | **L** | |
| | | | | P4 | 301393 | 661256 | 0.33333333 | 0.66666667 | 100464.33 | 440837.33 | 0.22789434 | **R** | |
| 8 | Argentina | 2 | 3 | P1 | 1102394 | 458590 | 0.66847826 | 0.33152174 | 736926.42 | 152032.55 | 4.84716201 | **L** | Left |
| | | | | P2 | 301725 | 180126 | 0.52542373 | 0.47457627 | 158533.47 | 85483.525 | 1.85455004 | **L** | |
| | | | | P3 | 1674419 | 1403836 | 0.5 | 0.5 | 837209.5 | 701918 | 1.19274545 | **L** | |
| 9 | Colombia | 2 | 4 | P1 | 266125 | 534171 | 0.5 | 0.5 | 133062.5 | 267085.5 | 0.49820189 | **R** | Mixed |
| | | | | P2 | 1265205 | 311493 | 0.84615385 | 0.15384615 | 1070558.1 | 47921.999 | 22.3395958 | **L** | |
| | | | | P3 | 453347 | 1381258 | 0.67032967 | 0.32967033 | 303891.94 | 455359.78 | 0.66736668 | **R** | |
| | | | | P4 | 544338 | 123766 | 0.74796748 | 0.25203252 | 407147.12 | 31193.057 | 13.0524919 | **L** | |
| 10 | Spain | 2 | 4 | P1 | 1552264 | 280078 | 0.88727273 | 0.11272727 | 1377281.5 | 31572.428 | 43.6229201 | **L** | Mixed |
| | | | | P2 | 814854 | 1065841 | 0.16574586 | 0.83425414 | 135058.68 | 889182.27 | 0.15189088 | **R** | |
| | | | | P3 | 638084 | 408021 | 0.5 | 0.5 | 319042 | 204010.5 | 1.56385088 | **L** | |
| | | | | P4 | 130570 | 1150131 | 0.00813008 | 0.99186992 | 1061.5447 | 1140780.3 | 0.00093054 | **R** | |
| 11 | Italy | 2 | 3 | P1 | 573638 | 2245835 | 0.89869281 | 0.10130719 | 515524.35 | 227519.23 | 2.26584952 | **L** | Left |
| | | | | P2 | 659634 | 675010 | 0.8 | 0.2 | 527707.2 | 135002 | 3.90888431 | **L** | |
| | | | | P3 | 90119 | 189963 | 0.79738562 | 0.20261438 | 71859.595 | 38489.235 | 1.867005 | **L** | |
| 12 | Iran | 2 | 3 | P1 | 1180537 | 192857 | 0.5 | 0.5 | 590268.5 | 96428.5 | 6.1213075 | **L** | Mixed |
| | | | | P2 | 1081078 | 705480 | 0.59333333 | 0.40666667 | 641439.61 | 286895.2 | 2.23579765 | **R** | |
| | | | | P3 | 666451 | 1121055 | 0.5 | 0.5 | 333225.5 | 560527.5 | 0.59448555 | **R** | |
| 13 | Germany | 2 | 2 | P1 | 1688712 | 689775 | 0.82335329 | 0.17664671 | 1390406.6 | 121846.48 | 11.4111342 | **L** | Left |
| | | | | P2 | 955070 | 560316 | 0.64893617 | 0.35106383 | 619779.47 | 196706.68 | 3.15077996 | **L** | |

| | | | | | | | | | | | | | |
|---|---|---|---|---|---|---|---|---|---|---|---|---|---|
| 14 | Indonesia | 2 | 4 | P1 | 230623 | 123080 | 0.74796748 | 0.25203252 | 172498.5 | 31020.163 | 5.56085107 | **L** | Mixed |
| | | | | P2 | 128795 | 204315 | 0.49180328 | 0.50819672 | 63341.803 | 103832.21 | 0.61004 | **R** | |
| | | | | P3 | 335116 | 590054 | 0.20529801 | 0.79470199 | 68798.649 | 468917.09 | 0.14671815 | **R** | |
| | | | | P4 | 2421433 | 172550 | 0.42990654 | 0.57009346 | 1040989.9 | 98369.626 | 10.5824321 | **L** | |
| 15 | Poland | 3 | 2 | P1 | 271217 | 1150654 | 0.25203252 | 0.74796748 | 68355.504 | 860651.77 | 0.07942295 | **R** | Mixed |
| | | | | P2 | 1278757 | 90797 | 0.41981132 | 0.58018868 | 536836.67 | 52679.392 | 10.1906391 | **L** | |
| 16 | Mexico | 2 | 3 | P1 | 580336 | 143656 | 0.80392157 | 0.19607843 | 466544.63 | 28167.843 | 16.5630229 | **L** | Mixed |
| | | | | P2 | 1121044 | 549482 | 0.4379562 | 0.5620438 | 490968.18 | 308832.95 | 1.58975322 | **L** | |
| | | | | P3 | 938668 | 603962 | 0.37908497 | 0.62091503 | 355834.93 | 375009.08 | 0.94887015 | **R** | |
| 17 | South Africa | 4 | 3 | P1 | 594358 | 98411 | 0.60130719 | 0.39869281 | 357391.74 | 39235.758 | 9.10882715 | **L** | Left |
| | | | | P2 | 728309 | 127449 | 0.50828729 | 0.49171271 | 370190.21 | 62668.293 | 5.90713723 | **L** | |
| | | | | P3 | 866244 | 330205 | 0.74796748 | 0.25203252 | 647922.34 | 83222.398 | 7.78543222 | **L** | |
| 18 | Ukraine | 3 | 2 | P1 | 1041743 | 174549 | 0.85581395 | 0.14418605 | 891538.19 | 25167.531 | 35.4241422 | **L** | Left |
| | | | | P2 | 862524 | 209339 | 0.41981132 | 0.58018868 | 362097.34 | 121456.12 | 2.98130177 | **L** | |
| | | | | P3 | 1273213 | 227330 | 0.74590164 | 0.25409836 | 949691.66 | 57764.18 | 16.4408403 | **L** | |
| 19 | Peru | 2 | 2 | P1 | 610190 | 315364 | 0.57476636 | 0.42523364 | 350716.69 | 134103.38 | 2.615271 | **L** | Mixed |
| | | | | P2 | 586277 | 600784 | 0.44160584 | 0.55839416 | 258903.35 | 335474.28 | 0.7717532 | **R** | |
| 20 | Netherlands | 2 | 7 | P1 | 26845 | 10971 | 0.19607843 | 0.80392157 | 5263.7255 | 8819.8235 | 0.59680621 | **R** | Mixed |
| | | | | P2 | 307157 | 174290 | 0.19607843 | 0.80392157 | 60226.862 | 140115.49 | 0.42983729 | **R** | |
| | | | | P3 | 276452 | 295182 | 0.19607843 | 0.80392157 | 54206.274 | 237303.18 | 0.22842625 | **R** | |
| | | | | P4 | 189242 | 230167 | 0.19607843 | 0.80392157 | 37106.274 | 185036.22 | 0.2005352 | **R** | |
| | | | | P5 | 153771 | 36003 | 0.25203252 | 0.74796748 | 38755.293 | 26929.073 | 1.43916177 | **L** | |
| | | | | P6 | 35744 | 180663 | 0.5 | 0.5 | 17872 | 90331.5 | 0.19784903 | **R** | |
| | | | | P7 | 779483 | 458444 | 0.74590164 | 0.25409836 | 581417.65 | 116489.87 | 4.99114346 | **L** | |
| 21 | Czechia | 2 | 2 | P1 | 264339 | 649672 | 0.20529801 | 0.79470199 | 54268.271 | 516295.63 | 0.10511085 | **R** | Right |
| | | | | P2 | 296852 | 655285 | 0.25203252 | 0.74796748 | 74816.358 | 490131.87 | 0.15264536 | **R** | |
| 22 | Iraq | 2 | 5 | P1 | 185825 | 128047 | 0.67391304 | 0.32608696 | 125229.89 | 41754.457 | 2.99919816 | **L** | Mixed |
| | | | | P2 | 189568 | 67087 | 0.25619835 | 0.74380165 | 48567.009 | 49899.421 | 0.97329804 | **R** | |

| | | | | | | | | | | | | | |
|---|---|---|---|---|---|---|---|---|---|---|---|---|---|
| | | | | P3 | 75853 | 155435 | 0.50847458 | 0.49152542 | 38569.322 | 76400.254 | 0.50483238 | **R** | |
| | | | | P4 | 214275 | 136153 | 0.01086957 | 0.98913043 | 2329.0761 | 134673.08 | 0.0172943 | **R** | |
| | | | | P5 | 686798 | 204986 | 0.20915033 | 0.79084967 | 143644.03 | 162113.11 | 0.88607285 | **R** | |
| 23 | Chile | 3 | 6 | P1 | 105848 | 288176 | 0.25203252 | 0.74796748 | 26677.138 | 215546.28 | 0.12376525 | **R** | Mixed |
| | | | | P2 | 51265 | 88752 | 0.32967033 | 0.67032967 | 16900.549 | 59493.099 | 0.2840758 | **R** | |
| | | | | P3 | 175366 | 97516 | 0.68888889 | 0.31111111 | 120807.69 | 30338.311 | 3.98201762 | **L** | |
| | | | | P4 | 170913 | 202707 | 0.50819672 | 0.49180328 | 86857.426 | 99691.967 | 0.87125802 | **R** | |
| | | | | P5 | 357657 | 59869 | 0.39869281 | 0.60130719 | 142595.27 | 35999.66 | 3.96101724 | **L** | |
| | | | | P6 | 40784 | 107843 | 0.32608696 | 0.67391304 | 13299.13 | 72676.804 | 0.18299003 | **R** | |
| 24 | Philippines | 3 | 3 | P1 | 202733 | 253245 | 0.42990654 | 0.57009346 | 87156.243 | 144373.32 | 0.60368663 | **R** | Mixed |
| | | | | P2 | 273224 | 841677 | 0.37037037 | 0.62962963 | 101194.07 | 529944.78 | 0.19095211 | **R** | |
| | | | | P3 | 1198311 | 52514 | 0.39869281 | 0.60130719 | 477757.98 | 31577.046 | 15.1299138 | **L** | |
| 25 | Canada | 3 | 5 | P1 | 47470 | 49313 | 0.32967033 | 0.67032967 | 15649.451 | 33055.967 | 0.47342286 | **R** | Mixed |
| | | | | P2 | 276679 | 204721 | 0.83152174 | 0.16847826 | 230064.6 | 34491.038 | 6.67027198 | **L** | |
| | | | | P3 | 196306 | 87508 | 0.49152542 | 0.50847458 | 96489.389 | 44495.594 | 2.16851561 | **L** | |
| | | | | P4 | 518571 | 48643 | 0.49456522 | 0.50543478 | 256467.18 | 24585.864 | 10.4314893 | **L** | |
| | | | | P5 | 193190 | 169220 | 0.30208333 | 0.69791667 | 58359.479 | 118101.46 | 0.49414698 | **R** | |
| 26 | Bangladesh | 3 | 5 | P1 | 98330 | 92178 | 0.49180328 | 0.50819672 | 48359.017 | 46844.557 | 1.03232946 | **L** | Mixed |
| | | | | P2 | 75335 | 94688 | 0.32608696 | 0.67391304 | 24565.761 | 63811.478 | 0.38497402 | **R** | |
| | | | | P3 | 105826 | 32706 | 0.50420168 | 0.49579832 | 53357.647 | 16215.58 | 3.29051736 | **L** | |
| | | | | P4 | 212916 | 41408 | 0.49593496 | 0.50406504 | 105592.49 | 20872.325 | 5.058971 | **L** | |
| | | | | P5 | 700078 | 83900 | 0.28504673 | 0.71495327 | 199554.94 | 59984.579 | 3.32677074 | **L** | |
| 27 | Malaysia | 1 | 3 | P1 | 183411 | 130541 | 0.60927152 | 0.39072848 | 111747.1 | 51006.087 | 2.19085812 | **L** | Mixed |
| | | | | P2 | 226857 | 179622 | 0.67032967 | 0.32967033 | 152068.98 | 59216.044 | 2.56803676 | **L** | |
| | | | | P3 | 361293 | 632982 | 0.5 | 0.5 | 180646.5 | 316491 | 0.57077926 | **R** | |
| 28 | Belgium | 2 | 3 | P1 | 105826 | 32706 | 0.64071856 | 0.35928144 | 67804.682 | 11750.659 | 5.77028774 | **L** | Mixed |
| | | | | P2 | 416454 | 342282 | 0.20261438 | 0.79738562 | 84379.569 | 272930.75 | 0.3091611 | **R** | |
| | | | | P3 | 680904 | 309746 | 0.32608696 | 0.67391304 | 222033.91 | 208741.87 | 1.06367694 | **L** | |

| | | | | | | | | | | | | | |
|---|---|---|---|---|---|---|---|---|---|---|---|---|---|
| 29 | Sweden | 1 | 3 | P1 | 369455 | 219930 | 0.75720165 | 0.24279835 | 279751.94 | 53398.641 | 5.23893361 | **L** | Mixed |
| | | | | P2 | 316295 | 126436 | 0.28773585 | 0.71226415 | 91009.41 | 90055.83 | 1.01058877 | **L** | |
| | | | | P3 | 26773 | 44699 | 0.19736842 | 0.80263158 | 5284.1447 | 35876.829 | 0.14728572 | **R** | |
| 30 | Romania | 2 | 3 | P1 | 424476 | 326632 | 0.575472 | 0.424528 | 244274.05 | 138664.43 | 1.76162014 | **L** | Left |
| | | | | P2 | 253271 | 22472 | 0.49593496 | 0.50406504 | 125605.94 | 11327.35 | 11.0887319 | **L** | |
| | | | | P3 | 564842 | 157695 | 0.79738562 | 0.20261438 | 450396.89 | 31951.275 | 14.0963669 | **L** | |
| 31 | Pakistan | 3 | 6 | P1 | 195356 | 99336 | 0.39869281 | 0.60130719 | 77887.033 | 59731.451 | 1.30395347 | **L** | Mixed |
| | | | | P2 | 87676 | 180883 | 0.40397351 | 0.59602649 | 35418.781 | 107811.06 | 0.32852642 | **R** | |
| | | | | P3 | 91566 | 152588 | 0.50819672 | 0.49180328 | 46533.541 | 75043.279 | 0.62008939 | **R** | |
| | | | | P4 | 97305 | 35584 | 0.50819672 | 0.49180328 | 49450.082 | 17500.328 | 2.82566601 | **L** | |
| | | | | P5 | 76429 | 128851 | 0.33695652 | 0.66304348 | 25753.25 | 85433.815 | 0.30144095 | **R** | |
| | | | | P6 | 82850 | 48323 | 0.24590164 | 0.75409836 | 20372.951 | 36440.295 | 0.55907755 | **R** | |
| 32 | Japan | 4 | 3 | P1 | 353932 | 84475 | 0.75720165 | 0.24279835 | 267997.89 | 20510.391 | 13.0664452 | **L** | Left |
| | | | | P2 | 272547 | 53117 | 0.5 | 0.5 | 136273.5 | 26558.5 | 5.13106915 | **L** | |
| | | | | P3 | 905489 | 26374 | 0.39869281 | 0.60130719 | 361011.95 | 15858.876 | 22.7640319 | **L** | |
| 33 | Portugal | 2 | 4 | P1 | 17602 | 17096 | 0.34065934 | 0.65934066 | 5996.2857 | 11272.088 | 0.53195874 | **R** | Mixed |
| | | | | P2 | 255920 | 115617 | 0.83695652 | 0.16304348 | 214193.91 | 18850.598 | 11.3627118 | **L** | |
| | | | | P3 | 306838 | 128577 | 0.20529801 | 0.79470199 | 62993.231 | 102180.4 | 0.61649037 | **R** | |
| | | | | P4 | 119538 | 69296 | 0.66304348 | 0.33695652 | 79258.891 | 23349.739 | 3.39442299 | **L** | |
| 34 | Israel | 3 | 3 | P1 | 241694 | 89780 | 0.66545455 | 0.33454545 | 160836.37 | 30035.491 | 5.35487749 | **L** | Mixed |
| | | | | P2 | 306244 | 196040 | 0.14553991 | 0.85446009 | 44570.723 | 167508.36 | 0.26608059 | **R** | |
| | | | | P3 | 442743 | 61442 | 0.32786885 | 0.67213115 | 145161.64 | 41297.082 | 3.51505802 | **L** | |
| 35 | Hungary | 3 | 3 | P1 | 241694 | 89780 | 0.66545455 | 0.33454545 | 160836.37 | 30035.491 | 5.35487749 | **L** | Mixed |
| | | | | P2 | 306244 | 198886 | 0.38410596 | 0.61589404 | 117630.15 | 122492.7 | 0.9603033 | **R** | |
| | | | | P3 | 293617 | 146586 | 0.9869281 | 0.0130719 | 289778.87 | 1916.1569 | 151.2292 | **L** | |
| 36 | Switzerland | 2 | 4 | P1 | 12981 | 1276 | 0.34065934 | 0.65934066 | 4422.0989 | 841.31868 | 5.25615202 | **L** | Left |
| | | | | P2 | 296210 | 227860 | 0.63374486 | 0.36625514 | 187721.56 | 83454.896 | 2.24937749 | **L** | |
| | | | | P3 | 105042 | 43030 | 0.40522876 | 0.59477124 | 42566.039 | 25593.007 | 1.66319026 | **L** | |

| | | | | | | | | | | | | | |
|---|---|---|---|---|---|---|---|---|---|---|---|---|---|
| | | | | P4 | 137359 | 32195 | 0.68131868 | 0.31868132 | 93585.253 | 10259.945 | 9.12141851 | **L** | |
| 37 | Nepal | 2 | 3 | P1 | 131283 | 103400 | 0.337017 | 0.662983 | 44244.603 | 68552.442 | 0.6454125 | **R** | Right |
| | | | | P2 | 45878 | 332536 | 0.33152174 | 0.66847826 | 15209.554 | 222293.09 | 0.06842118 | **R** | |
| | | | | P3 | 67258 | 65325 | 0.20261438 | 0.79738562 | 13627.438 | 52089.216 | 0.26161726 | **R** | |
| 38 | Austria | 2 | 3 | P1 | 272276 | 176984 | 0.668493 | 0.331507 | 182014.6 | 58671.435 | 3.10226945 | **L** | Mixed |
| | | | | P2 | 86789 | 104183 | 0.362983 | 0.637017 | 31502.932 | 66366.342 | 0.47468236 | **R** | |
| | | | | P3 | 509583 | 111775 | 0.79084967 | 0.20915033 | 403003.55 | 23377.778 | 17.238745 | **L** | |
| 39 | Kazakhstan | 2 | 3 | P1 | 68059 | 110829 | 0.163043 | 0.836957 | 11096.544 | 92759.107 | 0.11962754 | **R** | Mixed |
| | | | | P2 | 241739 | 39751 | 0.828729 | 0.171271 | 200336.12 | 6808.1935 | 29.4257381 | **L** | |
| | | | | P3 | 150783 | 230112 | 0.51612903 | 0.48387097 | 77823.484 | 111344.52 | 0.69894312 | **R** | |
| 40 | Tunisia | 1 | 4 | P1 | 41400 | 79327 | 0.33695652 | 0.66304348 | 13950 | 52597.25 | 0.26522299 | **R** | Mixed |
| | | | | P2 | 69745 | 24392 | 0.52542373 | 0.47457627 | 36645.678 | 11575.864 | 3.16569691 | **L** | |
| | | | | P3 | 20741 | 91456 | 0.336957 | 0.663043 | 6988.8251 | 60639.261 | 0.11525248 | **R** | |
| | | | | P4 | 244091 | 74469 | 0.66304348 | 0.33695652 | 161842.95 | 25092.815 | 6.44977235 | **L** | |
| 41 | Jordan | 3 | 2 | P1 | 207605 | 107425 | 0.49593496 | 0.50406504 | 102958.58 | 54149.187 | 1.90138731 | **L** | Left |
| | | | | P2 | 284722 | 139827 | 0.32596685 | 0.67403315 | 92809.934 | 94248.033 | 0.98474133 | **L** | |
| 42 | Serbia | 3 | 4 | P1 | 128484 | 219825 | 0.32608696 | 0.67391304 | 41896.957 | 148142.93 | 0.28281441 | **R** | Mixed |
| | | | | P2 | 205333 | 115966 | 0.39333333 | 0.60666667 | 80764.313 | 70352.707 | 1.14799156 | **R** | |
| | | | | P3 | 225427 | 200760 | 0.74796748 | 0.25203252 | 168612.07 | 50598.049 | 3.33238275 | **L** | |
| | | | | P4 | 112096 | 40353 | 0.49180328 | 0.50819672 | 55129.18 | 20507.262 | 2.68827596 | **L** | |
| 43 | UAE | 1 | 4 | P1 | 38439 | 36231 | 0.50819672 | 0.49180328 | 19534.574 | 17818.525 | 1.09630703 | **L** | Mixed |
| | | | | P2 | 134749 | 216627 | 0.41059603 | 0.58940397 | 55327.404 | 127680.81 | 0.4333259 | **R** | |
| | | | | P3 | 112671 | 47951 | 0.49593496 | 0.50406504 | 55877.488 | 24170.423 | 2.3118126 | **L** | |
| | | | | P4 | 55134 | 6049 | 0.24390244 | 0.75609756 | 13447.317 | 4573.6341 | 2.94018206 | **L** | |
| 44 | Morroco | 2 | 3 | P1 | 332014 | 127318 | 0.50205761 | 0.49794239 | 166690.16 | 63397.029 | 2.6293055 | **L** | Mixed |
| | | | | P2 | 15152 | 20112 | 0.32608696 | 0.67391304 | 4940.8696 | 13553.739 | 0.36453923 | **R** | |
| | | | | P3 | 329587 | 88969 | 0.39869281 | 0.60130719 | 131403.97 | 53497.699 | 2.45625455 | **L** | |
| 45 | Saudi Arabia | 2 | 4 | P1 | 168070 | 156459 | 0.33152174 | 0.66847826 | 55718.859 | 104589.44 | 0.53273886 | **R** | Mixed |

| | | | | | | | | | | | | | |
|---|---|---|---|---|---|---|---|---|---|---|---|---|---|
| | | | | P2 | 10078 | 5381 | 0.49180328 | 0.50819672 | 4956.3934 | 2734.6066 | 1.8124704 | **L** | |
| | | | | P3 | 5333 | 9309 | 0.65555556 | 0.34444444 | 3496.0778 | 3206.4333 | 1.09033228 | **L** | |
| | | | | P4 | 165935 | 6434 | 0.66393443 | 0.33606557 | 110169.96 | 2162.2459 | 50.9516327 | **L** | |
| 46 | Lebanon | 1 | 3 | P1 | 261418 | 73998 | 0.81456954 | 0.18543046 | 212943.14 | 13721.483 | 15.518959 | **L** | Mixed |
| | | | | P2 | 151528 | 18288 | 0.39869281 | 0.60130719 | 60413.124 | 10996.706 | 5.49374738 | **L** | |
| | | | | P3 | 57400 | 39758 | 0.25409836 | 0.74590164 | 14585.246 | 29655.557 | 0.49182167 | **R** | |
| 47 | Ecuador | 2 | 5 | P1 | 19969 | 60421 | 0.18584071 | 0.81415929 | 3711.0531 | 49192.319 | 0.07543969 | **R** | Mixed |
| | | | | P2 | 51692 | 75465 | 0.5 | 0.5 | 25846 | 37732.5 | 0.68497979 | **R** | |
| | | | | P3 | 38316 | 35327 | 0.68888889 | 0.31111111 | 26395.467 | 10990.622 | 2.40163533 | **L** | |
| | | | | P4 | 95707 | 76642 | 0.33152174 | 0.66847826 | 31728.951 | 51233.511 | 0.61930074 | **L** | |
| | | | | P5 | 28868 | 13829 | 0.53787879 | 0.46212121 | 15527.485 | 6390.6742 | 2.42970996 | **L** | |
| 48 | Bolivia | 2 | 3 | P1 | 83379 | 28110 | 0.40522876 | 0.59477124 | 33787.569 | 16719.02 | 2.02090609 | **L** | Mixed |
| | | | | P2 | 72127 | 55576 | 0.51239669 | 0.48760331 | 36957.636 | 27099.041 | 1.36379866 | **L** | |
| | | | | P3 | 167213 | 33882 | 0.59477124 | 0.40522876 | 99453.484 | 13729.961 | 7.24353734 | **L** | |
| 49 | Greece | 3 | 3 | P1 | 66020 | 51686 | 0.32608696 | 0.67391304 | 21528.261 | 34831.87 | 0.61806217 | **R** | Right |
| | | | | P2 | 106732 | 158767 | 0.39333333 | 0.60666667 | 41981.253 | 96318.647 | 0.43585801 | **R** | |
| | | | | P3 | 70848 | 94660 | 0.5 | 0.5 | 35424 | 47330 | 0.74844707 | **R** | |
| 50 | Paraguay | 1 | 3 | P1 | 92469 | 26247 | 0.81456954 | 0.18543046 | 75322.43 | 4866.9934 | 15.4761728 | **L** | Mixed |
| | | | | P2 | 119603 | 76307 | 0.66304348 | 0.33695652 | 79301.989 | 25712.141 | 3.08422345 | **L** | |
| | | | | P3 | 67898 | 35246 | 0.32608696 | 0.67391304 | 22140.652 | 23752.739 | 0.93213048 | **R** | |
| 51 | Belarus | 2 | 4 | P1 | 38165 | 57637 | 0.49180328 | 0.50819672 | 18769.672 | 29290.934 | 0.64080141 | **R** | Mixed |
| | | | | P2 | 93022 | 34501 | 0.65555556 | 0.34444444 | 60981.089 | 11883.678 | 5.13149969 | **L** | |
| | | | | P3 | 72632 | 22750 | 0.5 | 0.5 | 36316 | 11375 | 3.19261538 | **L** | |
| | | | | P4 | 51601 | 92046 | 0.25409836 | 0.74590164 | 13111.73 | 68657.262 | 0.19097367 | **R** | |
| 52 | Bulgaria | 3 | 3 | P1 | 92456 | 73448 | 0.32608696 | 0.67391304 | 30148.696 | 49497.565 | 0.60909452 | **R** | Mixed |
| | | | | P2 | 185632 | 17449 | 0.39333333 | 0.60666667 | 73015.253 | 10585.727 | 6.89751924 | **L** | |
| | | | | P3 | 180663 | 138190 | 0.60927152 | 0.39072848 | 110072.82 | 53994.768 | 2.03858309 | **L** | |
| 53 | Panama | 2 | 3 | P1 | 47339 | 19613 | 0.67391304 | 0.32608696 | 31902.37 | 6395.5435 | 4.9882187 | **L** | Left |

| | | | | | | | | | | | | | |
|---|---|---|---|---|---|---|---|---|---|---|---|---|---|
| | | | | P2 | 207784 | 44197 | 0.60666667 | 0.39333333 | 126055.63 | 17384.153 | 7.2511801 | **L** | |
| | | | | P3 | 39202 | 31877 | 0.70754717 | 0.29245283 | 27737.264 | 9322.5189 | 2.97529719 | **L** | |
| 54 | Slovakia | 2 | 2 | P1 | 121879 | 128540 | 0.40397351 | 0.59602649 | 49235.887 | 76613.245 | 0.64265503 | **R** | Right |
| | | | | P2 | 53102 | 33738 | 0.20261438 | 0.79738562 | 10759.229 | 26902.196 | 0.39993868 | **R** | |
| 55 | Costa Rica | 2 | 4 | P1 | 34473 | 34211 | 0.49180328 | 0.50819672 | 16953.934 | 17385.918 | 0.97515325 | **R** | Mixed |
| | | | | P2 | 59350 | 35020 | 0.50833333 | 0.49166667 | 30169.583 | 17218.167 | 1.75219487 | **L** | |
| | | | | P3 | 114645 | 48952 | 0.60130719 | 0.39869281 | 68936.863 | 19516.81 | 3.53217873 | **L** | |
| | | | | P4 | 164247 | 38371 | 0.39869281 | 0.60130719 | 65484.098 | 23072.758 | 2.83815648 | **L** | |
| 56 | Georgia | 2 | 4 | P1 | 188484 | 54341 | 0.40397351 | 0.59602649 | 76142.543 | 32388.675 | 2.35090018 | **L** | Mixed |
| | | | | P2 | 28549 | 55768 | 0.24590164 | 0.75409836 | 7020.2459 | 42054.557 | 0.16693187 | **R** | |
| | | | | P3 | 183733 | 63201 | 0.68478261 | 0.31521739 | 125817.16 | 19922.054 | 6.31547133 | **L** | |
| | | | | P4 | 106235 | 211297 | 0.31372549 | 0.68627451 | 33328.627 | 145007.75 | 0.22984033 | **R** | |
| 57 | Kuwait | 2 | 4 | P1 | 20073 | 20744 | 0.79738562 | 0.20261438 | 16005.922 | 4203.0327 | 3.80818394 | **L** | Left |
| | | | | P2 | 16709 | 7949 | 0.49180328 | 0.50819672 | 8217.541 | 4039.6557 | 2.03421814 | **L** | |
| | | | | P3 | 81519 | 41888 | 0.75 | 0.25 | 61139.25 | 10472 | 5.83835466 | **L** | |
| | | | | P4 | 123840 | 12029 | 0.74796748 | 0.25203252 | 92628.293 | 3031.6992 | 30.5532597 | **L** | |
| 58 | Thailand | 1 | 4 | P1 | 130929 | 99509 | 0.8 | 0.2 | 104743.2 | 19901.8 | 5.26300134 | **L** | Mixed |
| | | | | P2 | 337986 | 607442 | 0.50819672 | 0.49180328 | 171763.38 | 298741.97 | 0.57495563 | **R** | |
| | | | | P3 | 398746 | 308549 | 0.48387097 | 0.51612903 | 192941.61 | 159251.1 | 1.21155594 | **L** | |
| | | | | P4 | 203848 | 101415 | 0.47540984 | 0.52459016 | 96911.344 | 53201.311 | 1.8215969 | **L** | |
| 59 | Uruguay | 1 | 2 | P1 | 311356 | 15584 | 0.82872928 | 0.17127072 | 258029.83 | 2669.0829 | 96.6735941 | **R** | Mixed |
| | | | | P2 | 8965 | 5796 | 0.98901099 | 0.01098901 | 8866.4835 | 63.692308 | 139.208075 | **L** | |
| 60 | Croatia | 3 | 3 | P1 | 161521 | 32136 | 0.50833333 | 0.49166667 | 82106.508 | 15800.2 | 5.19654867 | **L** | Left |
| | | | | P2 | 89210 | 27689 | 0.33152174 | 0.66847826 | 29575.054 | 18509.495 | 1.59783155 | **L** | |
| | | | | P3 | 244590 | 95515 | 0.59477124 | 0.40522876 | 145475.1 | 38705.425 | 3.7585196 | **L** | |
| 61 | Azerbaijan | 3 | 3 | P1 | 163431 | 15837 | 0.50833333 | 0.49166667 | 83077.425 | 7786.525 | 10.669384 | **L** | Mixed |
| | | | | P2 | 27176 | 74334 | 0.20261438 | 0.79738562 | 5506.2484 | 59272.863 | 0.09289662 | **R** | |
| | | | | P3 | 147855 | 131940 | 0.33152174 | 0.66847826 | 49017.147 | 88199.022 | 0.55575613 | **R** | |

| | | | | | | | | | | | | | |
|---|---|---|---|---|---|---|---|---|---|---|---|---|---|
| 62 | Dominican Republic | 4 | 3 | P1 | 87042 | 38667 | 0.75720165 | 0.24279835 | 65908.346 | 9388.284 | 7.02027613 | **L** | Mixed |
| | | | | P2 | 40059 | 57387 | 0.39869281 | 0.60130719 | 15971.235 | 34507.216 | 0.46283755 | **R** | |
| | | | | P3 | 57078 | 9019 | 0.63114754 | 0.36885246 | 36024.639 | 3326.6803 | 10.8290054 | **L** | |
| 63 | Guatemala | 2 | 3 | P1 | 51565 | 15038 | 0.86792453 | 0.13207547 | 44754.528 | 1986.1509 | 22.5332966 | **L** | Mixed |
| | | | | P2 | 53129 | 26746 | 0.49593496 | 0.50406504 | 26348.528 | 13481.724 | 1.95438872 | **L** | |
| | | | | P3 | 215860 | 156398 | 0.28504673 | 0.71495327 | 61530.187 | 111817.26 | 0.55027449 | **R** | |
| 64 | Palestine | 3 | 5 | P1 | 52357 | 20958 | 0.48387097 | 0.51612903 | 25334.032 | 10817.032 | 2.34205017 | **L** | Mixed |
| | | | | P2 | 24650 | 58741 | 0.47457627 | 0.52542373 | 11698.305 | 30863.915 | 0.37902855 | **R** | |
| | | | | P3 | 54109 | 20276 | 0.24590164 | 0.75409836 | 13305.492 | 15290.098 | 0.87020315 | **R** | |
| | | | | P4 | 25544 | 61434 | 0.01639344 | 0.98360656 | 418.7541 | 60426.885 | 0.00692993 | **R** | |
| | | | | P5 | 49281 | 7056 | 0.01408451 | 0.98591549 | 694.09859 | 6956.6197 | 0.09977527 | **R** | |
| 65 | Denmark | 1 | 3 | P1 | 117253 | 47767 | 0.19607843 | 0.80392157 | 22990.784 | 38400.922 | 0.59870397 | **L** | Left |
| | | | | P2 | 19412 | 19954 | 0.50819672 | 0.49180328 | 9865.1148 | 9813.4426 | 1.00526544 | **L** | |
| | | | | P3 | 43254 | 23406 | 0.49593496 | 0.50406504 | 21451.171 | 11798.146 | 1.81818144 | **L** | |
| 66 | Venezuela | 2 | 4 | P1 | 45285 | 10381 | 0.67032967 | 0.32967033 | 30355.879 | 3422.3077 | 8.87000289 | **L** | Mixed |
| | | | | P2 | 24533 | 12189 | 0.68888889 | 0.31111111 | 16900.511 | 3792.1333 | 4.45672914 | **L** | |
| | | | | P3 | 21381 | 37186 | 0.33695652 | 0.66304348 | 7204.4674 | 24655.935 | 0.29220013 | **R** | |
| | | | | P4 | 171285 | 75015 | 0.62704918 | 0.37295082 | 107404.12 | 27976.906 | 3.8390278 | **L** | |
| 67 | Oman | 2 | 4 | P1 | 67722 | 6563 | 0.66304348 | 0.33695652 | 44902.63 | 2211.4457 | 20.3046502 | **L** | Mixed |
| | | | | P2 | 28712 | 14433 | 0.5 | 0.5 | 14356 | 7216.5 | 1.98933001 | **L** | |
| | | | | P3 | 64386 | 23971 | 0.79470199 | 0.20529801 | 51167.682 | 4921.1987 | 10.3974022 | **L** | |
| | | | | P4 | 51321 | 27312 | 0.32608696 | 0.67391304 | 16735.109 | 18405.913 | 0.90922459 | **R** | |
| 68 | Egypt | 3 | 5 | P1 | 69093 | 9120 | 0.5 | 0.5 | 34546.5 | 4560 | 7.57598684 | **L** | Mixed |
| | | | | P2 | 34864 | 64069 | 0.50549451 | 0.49450549 | 17623.56 | 31682.473 | 0.55625584 | **R** | |
| | | | | P3 | 25421 | 35098 | 0.49180328 | 0.50819672 | 12502.131 | 17836.689 | 0.70092221 | **R** | |
| | | | | P4 | 18632 | 2980 | 0.32608696 | 0.67391304 | 6075.6522 | 2008.2609 | 3.02533016 | **L** | |
| | | | | P5 | 20262 | 26493 | 0.65217391 | 0.34782609 | 13214.348 | 9214.9565 | 1.43401087 | **L** | |

| | | | | | | | | | | | | | |
|---|---|---|---|---|---|---|---|---|---|---|---|---|---|
| 69 | Ireland | 1 | 3 | P1 | 35983 | 11088 | 0.85981308 | 0.14018692 | 30938.654 | 1554.3925 | 19.9040164 | **L** | Mixed |
| | | | | P2 | 124003 | 65496 | 0.34065934 | 0.65934066 | 42242.78 | 43184.176 | 0.97820045 | **R** | |
| | | | | P3 | 38933 | 51471 | 0.66304348 | 0.33695652 | 25814.272 | 17343.489 | 1.48841283 | **L** | |
| 70 | Sri Lanka | 1 | 2 | P1 | 78218 | 122448 | 0.33695652 | 0.66304348 | 26356.065 | 81188.348 | 0.32462867 | **R** | Right |
| | | | | P2 | 131490 | 277371 | 0.25409836 | 0.74590164 | 33411.393 | 206891.48 | 0.16149236 | **R** | |
| 71 | Lithuania | 2 | 3 | P1 | 127140 | 74164 | 0.40397351 | 0.59602649 | 51361.192 | 44203.709 | 1.16192043 | **L** | Mixed |
| | | | | P2 | 58264 | 8435 | 0.39869281 | 0.60130719 | 23229.438 | 5072.0261 | 4.57991289 | **L** | |
| | | | | P3 | 125897 | 107177 | 0.39869281 | 0.60130719 | 50194.229 | 64446.301 | 0.77885353 | **R** | |
| 72 | Honduras | 1 | 4 | P1 | 15886 | 19988 | 0.74796748 | 0.25203252 | 11882.211 | 5037.626 | 2.35869263 | **L** | Mixed |
| | | | | P2 | 24939 | 67216 | 0.60927152 | 0.39072848 | 15194.623 | 26263.205 | 0.57855172 | **R** | |
| | | | | P3 | 23290 | 25894 | 0.32608696 | 0.67391304 | 7594.5652 | 17450.304 | 0.43521105 | **R** | |
| | | | | P4 | 100530 | 40645 | 0.28504673 | 0.71495327 | 28655.748 | 29059.276 | 0.98611362 | **R** | |
| 73 | Ethiopia | 2 | 2 | P1 | 27013 | 72133 | 0.13475177 | 0.86524823 | 3640.0496 | 62412.95 | 0.05832202 | **R** | Right |
| | | | | P2 | 82325 | 69585 | 0.4245283 | 0.5754717 | 34949.292 | 40044.198 | 0.87276794 | **R** | |
| 74 | Bahrain | 2 | 2 | P1 | 96086 | 32009 | 0.5 | 0.5 | 48043 | 16004.5 | 3.00184323 | **L** | Left |
| | | | | P2 | 2517 | 2633 | 0.66304348 | 0.33695652 | 1668.8804 | 887.20652 | 1.88105068 | **L** | |
| 75 | Cuba | 1 | 3 | P1 | 2095 | 1241 | 0.65555556 | 0.34444444 | 1373.3889 | 427.45556 | 3.21293962 | **L** | Mixed |
| | | | | P2 | 13786 | 492832 | 0.7398374 | 0.2601626 | 10199.398 | 128216.46 | 0.07954828 | **R** | |
| | | | | P3 | 74573 | 13242 | 0.98913043 | 0.01086957 | 73762.424 | 143.93478 | 512.471152 | **L** | |
| 76 | Slovenia | 2 | 3 | P1 | 132166 | 49129 | 0.60927152 | 0.39072848 | 80524.98 | 19196.099 | 4.19486161 | **L** | Mixed |
| | | | | P2 | 24690 | 18923 | 0.19607843 | 0.80392157 | 4841.1765 | 15212.608 | 0.31823449 | **R** | |
| | | | | P3 | 161683 | 39537 | 0.59477124 | 0.40522876 | 96164.399 | 16021.529 | 6.00219843 | **L** | |
| 77 | Moldova | 2 | 3 | P1 | 91776 | 14986 | 0.74796748 | 0.25203252 | 68645.463 | 3776.9593 | 18.1747954 | **L** | Left |
| | | | | P2 | 70437 | 26493 | 0.32596685 | 0.67403315 | 22960.127 | 17857.16 | 1.28576587 | **L** | |
| | | | | P3 | 81034 | 37590 | 0.5 | 0.5 | 40517 | 18795 | 2.15573291 | **L** | |
| 78 | Armenia | 4 | 2 | P1 | 32247 | 82245 | 0.13043478 | 0.86956522 | 4206.1304 | 71517.391 | 0.05881269 | **R** | Right |
| | | | | P2 | 20581 | 32456 | 0.16847826 | 0.83152174 | 3467.4511 | 26987.87 | 0.12848184 | **R** | |
| 79 | Qatar | 2 | 4 | P1 | 39178 | 29672 | 0.24590164 | 0.75409836 | 9633.9344 | 22375.607 | 0.43055523 | **R** | Mixed |

| | | | | | | | | | | | | | |
|---|---|---|---|---|---|---|---|---|---|---|---|---|---|
| | | | | P2 | 6796 | 11278 | 0.33695652 | 0.66304348 | 2289.9565 | 7477.8043 | 0.30623381 | **R** | |
| | | | | P3 | 73624 | 8781 | 0.62139918 | 0.37860082 | 45749.893 | 3324.4938 | 13.7614613 | **L** | |
| | | | | P4 | 10404 | 6804 | 0.20261438 | 0.79738562 | 2108 | 5425.4118 | 0.38854194 | **R** | |
| 80 | Libya | 2 | 3 | P1 | 26570 | 39182 | 0.33695652 | 0.66304348 | 8952.9348 | 25979.37 | 0.34461709 | **R** | Mixed |
| | | | | P2 | 77231 | 15966 | 0.66298343 | 0.33701657 | 51202.873 | 5380.8066 | 9.51583591 | **L** | |
| | | | | P3 | 55640 | 59858 | 0.5 | 0.5 | 27820 | 29929 | 0.92953323 | **R** | |
| 81 | Afghanistan | 2 | **2** | P1 | 91500 | 6697 | 0.85377358 | 0.14622642 | 78120.283 | 979.2783 | 79.7733217 | **L** | Left |
| | | | | P2 | 91440 | 10883 | 0.69281046 | 0.30718954 | 63350.588 | 3343.1438 | 18.9494058 | **L** | |
| 82 | Albania | 2 | 3 | P1 | 17990 | 7364 | 0.25619835 | 0.74380165 | 4609.0083 | 5477.3554 | 0.84146599 | **R** | Mixed |
| | | | | P2 | 13866 | 38913 | 0.51219512 | 0.48780488 | 7102.0976 | 18981.951 | 0.37415003 | **R** | |
| | | | | P3 | 14645 | 8954 | 0.49180328 | 0.50819672 | 7202.459 | 4550.3934 | 1.58282116 | **L** | |
| 83 | Algeria | 3 | 4 | P1 | 16411 | 7729 | 0.5 | 0.5 | 8205.5 | 3864.5 | 2.12330185 | **L** | Mixed |
| | | | | P2 | 5753 | 4100 | 0.47457627 | 0.52542373 | 2730.2373 | 2154.2373 | 1.26738002 | **L** | |
| | | | | P3 | 4916 | 17518 | 0.32967033 | 0.67032967 | 1620.6593 | 11742.835 | 0.13801261 | **R** | |
| | | | | P4 | 31766 | 24688 | 0.5 | 0.5 | 15883 | 12344 | 1.28669799 | **L** | |
| 84 | Belize | 2 | 2 | P1 | 6814 | 141 | 0.92907801 | 0.07092199 | 6330.7376 | 10 | 633.073759 | **L** | Left |
| | | | | P2 | 13989 | 5269 | 0.31304348 | 0.68695652 | 4379.1652 | 3619.5739 | 1.20985655 | **L** | |
| 85 | Bosnia and Herzegovina | 3 | 4 | P1 | 37811 | 43078 | 0.25 | 0.75 | 9452.75 | 32308.5 | 0.2925778 | **L** | Mixed |
| | | | | P2 | 38647 | 36029 | 0.85620915 | 0.14379085 | 33089.915 | 5180.6405 | 6.38722469 | **L** | |
| | | | | P3 | 29120 | 17983 | 0.66304348 | 0.33695652 | 19307.826 | 6059.4891 | 3.18637853 | **R** | |
| | | | | P4 | 22307 | 14611 | 0.49180328 | 0.50819672 | 10970.656 | 7425.2623 | 1.4774772 | **L** | |
| 86 | Botswana | 1 | 2 | P1 | 33206 | 7086 | 0.83425414 | 0.16574586 | 27702.243 | 1174.4751 | 23.5869132 | **L** | Left |
| | | | | P2 | 109993 | 29667 | 0.66847826 | 0.33152174 | 73527.929 | 9835.2554 | 7.47595523 | **L** | |
| 87 | Cambodia | 1 | 1 | P1 | 26858 | 15812 | 0.5 | 0.5 | 13429 | 7906 | 1.69858335 | **L** | Left |
| 88 | Cameroon | 1 | 3 | P1 | 4663 | 9201 | 0.11510791 | 0.88489209 | 536.7482 | 8141.8921 | 0.06592426 | **R** | Mixed |
| | | | | P2 | 2652 | 1832 | 0.47540984 | 0.52459016 | 1260.7869 | 961.04918 | 1.31188592 | **L** | |
| | | | | P3 | 45973 | 9814 | 0.56603774 | 0.43396226 | 26022.453 | 4258.9057 | 6.11012662 | **L** | |

| | | | | | | | | | | | | | |
|---|---|---|---|---|---|---|---|---|---|---|---|---|---|
| 89 | China | 1 | 1 | P1 | 69471 | 2304 | 0.11764706 | 0.88235294 | 8173.0588 | 2032.9412 | 4.0203125 | **L** | Left |
| 90 | **Cyprus** | 3 | 0 | | | | | | | | | | |
| 91 | El Salvador | 2 | 1 | P1 | 17728 | 17393 | 0.57009346 | 0.42990654 | 10106.617 | 7477.3645 | 1.35162822 | **L** | Left |
| 92 | Equatorial Guinea | 1 | 2 | P1 | 2820 | 267 | 0.24590164 | 0.75409836 | 693.44262 | 201.34426 | 3.44406448 | **L** | Left |
| | | | | P2 | 3441 | 351 | 0.69736842 | 0.30263158 | 2399.6447 | 106.22368 | 22.5904868 | **L** | |
| 93 | Estonia | 2 | 2 | P1 | 62216 | 24640 | 0.33149171 | 0.66850829 | 20624.088 | 16472.044 | 1.25206612 | **L** | Left |
| | | | | P2 | 91520 | 16373 | 0.66847826 | 0.33152174 | 61179.13 | 5428.0054 | 11.271015 | **L** | |
| 94 | Eswatini | 3 | 1 | P1 | 1082 | 137 | 0.40425532 | 0.59574468 | 437.40426 | 81.617021 | 5.35922836 | **L** | Left |
| 95 | **Fiji** | 1 | **0** | | | | | | | | | | |
| 96 | Ghana | 3 | 3 | P1 | 17760 | 12554 | 0.25203252 | 0.74796748 | 4476.0976 | 9389.9837 | 0.47668853 | **R** | Mixed |
| | | | | P2 | 3612 | 3104 | 0.49180328 | 0.50819672 | 1776.3934 | 1577.4426 | 1.12612238 | **L** | |
| | | | | P3 | 12239 | 26888 | 0.17032967 | 0.82967033 | 2084.6648 | 22308.176 | 0.09344847 | **R** | |
| 97 | Kenya | 4 | 3 | P1 | 37474 | 17155 | 0.32608696 | 0.67391304 | 12219.783 | 11560.978 | 1.05698517 | **L** | Mixed |
| | | | | P2 | 33285 | 36677 | 0.65555556 | 0.34444444 | 21820.167 | 12633.189 | 1.72720972 | **R** | |
| | | | | P3 | 65128 | 19225 | 0.66666667 | 0.33333333 | 43418.667 | 6408.3333 | 6.7753446 | **L** | |
| 98 | Kyrgyzstan | 3 | 3 | P1 | 22704 | 10864 | 0.18666667 | 0.81333333 | 4238.08 | 8836.0533 | 0.47963495 | **R** | Mixed |
| | | | | P2 | 26509 | 13073 | 0.40397351 | 0.59602649 | 10708.934 | 7791.8543 | 1.37437552 | **L** | |
| | | | | P3 | 38752 | 37889 | 0.66304348 | 0.33695652 | 25694.261 | 12766.946 | 2.01256131 | **L** | |
| 99 | Latvia | 2 | 3 | P1 | 14432 | 61459 | 0.1509434 | 0.8490566 | 2178.4151 | 52182.17 | 0.04174635 | **R** | Mixed |
| | | | | P2 | 30836 | 5664 | 0.39215686 | 0.60784314 | 12092.549 | 3442.8235 | 3.51239293 | **L** | |
| | | | | P3 | 80276 | 55132 | 0.39215686 | 0.60784314 | 31480.784 | 33511.608 | 0.9393994 | **R** | |
| 100 | Luxembourg | 1 | 3 | P1 | 4283 | 29281 | 0.32608696 | 0.67391304 | 1396.6304 | 19732.848 | 0.07077693 | **R** | Right |
| | | | | P2 | 11254 | 9145 | 0.32596685 | 0.67403315 | 3668.4309 | 6164.0331 | 0.59513485 | **R** | |
| | | | | P3 | 1890 | 2459 | 0.5 | 0.5 | 945 | 1229.5 | 0.76860512 | **R** | |
| 101 | Madagascar | 2 | 2 | P1 | 16859 | 5915 | 0.24590164 | 0.75409836 | 4145.6557 | 4460.4918 | 0.92941674 | **R** | Mixed |
| | | | | P2 | 728 | 704 | 0.49180328 | 0.50819672 | 358.03279 | 357.77049 | 1.00073314 | **L** | |
| 102 | Malawi | 2 | 2 | P1 | 13972 | 10375 | 0.12408759 | 0.87591241 | 1733.7518 | 9087.5912 | 0.19078233 | **R** | Mixed |

| | | | | | | | | | | | | | |
|---|---|---|---|---|---|---|---|---|---|---|---|---|---|
| | | | | P2 | 18009 | 8147 | 0.70114943 | 0.29885057 | 12627 | 2434.7356 | 5.18618935 | **L** | |
| 103 | Maldives | 1 | 1 | P1 | 29672 | 13036 | 0.20175439 | 0.79824561 | 5986.4561 | 10405.93 | 0.57529276 | **R** | Right |
| 104 | Mongolia | 1 | 2 | P1 | 27687 | 21154 | 0.46551724 | 0.53448276 | 12888.776 | 11306.448 | 1.13994913 | **L** | Left |
| | | | | P2 | 105716 | 49665 | 0.65217391 | 0.34782609 | 68945.217 | 17274.783 | 3.99109031 | **L** | |
| 105 | Mozambique | 2 | 1 | P1 | 31080 | 11445 | 0.09090909 | 0.90909091 | 2825.4545 | 10404.545 | 0.27155963 | **R** | Right |
| 106 | Myanmar | 2 | 2 | P1 | 111257 | 18187 | 0.20529801 | 0.79470199 | 22840.841 | 14453.245 | 1.58032615 | **L** | Left |
| | | | | P2 | 155556 | 100097 | 0.79738562 | 0.20261438 | 124038.12 | 20281.092 | 6.11594882 | **L** | |
| 107 | Namibia | 1 | 1 | P1 | 29588 | 29005 | 0.12068966 | 0.87931034 | 3570.9655 | 25504.397 | 0.14001372 | **R** | Right |
| 108 | Nigeria | 3 | 2 | P1 | 80595 | 10861 | 0.23333333 | 0.76666667 | 18805.5 | 8326.7667 | 2.25843965 | **L** | Left |
| | | | | P2 | 25913 | 21787 | 0.49726776 | 0.50273224 | 12885.699 | 10953.027 | 1.17645095 | **L** | |
| 109 | North Macedonia | 2 | 4 | P1 | 52841 | 19458 | 0.50833333 | 0.49166667 | 26860.842 | 9566.85 | 2.80769968 | **L** | Left |
| | | 1 | | P2 | 49580 | 3317 | 0.39869281 | 0.60130719 | 19767.19 | 1994.5359 | 9.91067098 | **L** | |
| | | | | P3 | 20810 | 26058 | 0.5754717 | 0.4245283 | 11975.566 | 11062.358 | 1.08255089 | **L** | |
| | | | | P4 | 13033 | 8549 | 0.90196078 | 0.09803922 | 11755.255 | 838.13725 | 14.0254533 | **L** | |
| 110 | Norway | 1 | 4 | P1 | 7716 | 13417 | 0.31111111 | 0.68888889 | 2400.5333 | 9242.8222 | 0.25971865 | **R** | Mixed |
| | | | | P2 | 13399 | 8040 | 0.52542373 | 0.47457627 | 7040.1525 | 3815.5932 | 1.84510039 | **L** | |
| | | | | P3 | 25073 | 35237 | 0.20261438 | 0.79738562 | 5080.1503 | 28097.477 | 0.1808045 | **R** | |
| | | | | P4 | 51804 | 17849 | 0.33695652 | 0.66304348 | 17455.696 | 11834.663 | 1.47496347 | **L** | |
| 111 | Rwanda | 2 | 1 | P1 | 9890 | 2832 | 0.98901099 | 0.01098901 | 9781.3187 | 31.120879 | 314.300847 | **L** | |
| 112 | South Korea | 1 | 4 | P1 | 36772 | 25131 | 0.20529801 | 0.79470199 | 7549.2185 | 19971.656 | 0.37799663 | **R** | Mixed |
| | | | | P2 | 18995 | 18165 | 0.24590164 | 0.75409836 | 4670.9016 | 13698.197 | 0.34098661 | **R** | |
| | | | | P3 | 58988 | 53658 | 0.74796748 | 0.25203252 | 44121.106 | 13523.561 | 3.26253609 | **L** | |
| | | | | P4 | 60328 | 52613 | 0.49180328 | 0.50819672 | 29669.508 | 26737.754 | 1.10964848 | **L** | |
| 113 | Senegal | 2 | 0 | | | | | | | | | | |
| 114 | Seychelles | 1 | 0 | | | | | | | | | | |
| 115 | Singapore | 2 | 5 | P1 | 9581 | 36036 | 0.10784314 | 0.89215686 | 1033.2451 | 32149.765 | 0.0321385 | **R** | Mixed |
| | | | | P2 | 4607 | 1203 | 0.34408602 | 0.65591398 | 1585.2043 | 789.06452 | 2.00896665 | **L** | |

| | | | | | | | | | | | | | |
|---|---|---|---|---|---|---|---|---|---|---|---|---|---|
| | | | | P3 | 1521 | 845 | 0.60927152 | 0.39072848 | 926.70199 | 330.16556 | 2.80677966 | **L** | |
| | | | | P4 | 764 | 1434 | 0.32967033 | 0.67032967 | 251.86813 | 961.25275 | 0.26202071 | **R** | |
| | | | | P5 | 2402 | 2639 | 0.48387097 | 0.51612903 | 1162.2581 | 1362.0645 | 0.85330618 | **R** | |
| 116 | Sudan | 2 | 2 | P1 | 3597 | 3194 | 0.45856354 | 0.54143646 | 1649.453 | 1729.3481 | 0.95380049 | **L** | Mixed |
| | | | | P2 | 571 | 574 | 0.43396226 | 0.56603774 | 247.79245 | 324.90566 | 0.7626597 | **R** | |
| 117 | Uganda | 2 | 2 | P1 | 12016 | 5651 | 0.39333333 | 0.60666667 | 4726.2933 | 3428.2733 | 1.37862209 | **R** | Mixed |
| | | | | P2 | 39110 | 13950 | 0.74485597 | 0.25514403 | 29131.317 | 3559.2593 | 8.18465719 | **L** | |
| 118 | Vietnam | 1 | 2 | P1 | 312036 | 459026 | 0.67391304 | 0.32608696 | 210285.13 | 149682.39 | 1.40487554 | **L** | Mixed |
| | | 2 | | P2 | 316960 | 456792 | 0.49180328 | 0.50819672 | 155881.97 | 232140.2 | 0.67149925 | **R** | |
| 119 | Zambia | 2 | 2 | P1 | 29920 | 37369 | 0.32608696 | 0.67391304 | 9756.5217 | 25183.457 | 0.3874179 | **R** | Mixed |
| | | | | P2 | 63362 | 40868 | 0.70754717 | 0.29245283 | 44831.604 | 11951.962 | 3.75098271 | **L** | |
| 120 | Zimbabwe | 2 | 2 | P1 | 17559 | 3494 | 0.31395349 | 0.68604651 | 5512.7093 | 2397.0465 | 2.29979238 | **L** | Left |
| | | | | P2 | 69899 | 98688 | 0.5 | 0.5 | 34949.5 | 49344 | 0.70828267 | **L** | |
